# Comparing methods to estimate time-varying reproduction numbers using genomic and epidemiological data

**DOI:** 10.1101/2025.09.25.25336592

**Authors:** Elisha B. Are, Siavash Riazi, Niloufar Saeidi Mobarakeh, Jessica E. Stockdale, Caroline Colijn

## Abstract

Estimating the time-varying reproduction number ℛ_*t*_ during an epidemic is important. ℛ_*t*_ indicates whether an epidemic is growing or declining and can aid in assessing the impact of interventions. Recent advances have enhanced methods for estimating ℛ_*t*_ and other epidemiological parameters from surveillance and genomic data independently. The Birth-Death Skyline (BDSKY) in BEAST 2.5 and EpiEstim are two common methods used to estimate ℛ_*t*_ from these data sources. We introduce an outbreak simulation platform that generates pathogen sequence data and epidemiological linelists. We use this platform to to assess ℛ_*t*_ estimation methods’ accuracy under various sampling scenarios similar to what was observed during past epidemics. We identified biases and determined appropriate scenarios for improving the accuracy of ℛ_*t*_ estimation approaches based on multiple outbreak simulations. When data becomes sparse and unreliable, genomic sequence data provide reasonable ℛ_*t*_ estimates even when sampling is not uniform.

## Introduction

Accurately estimating the time-varying reproduction number (ℛ_*t*_, the expected number of infections an infected individual generates throughout the period of infectiousness, at a given time period) in emerging epidemics can be challenging due to limited knowledge about the pathogen and constraints on resources for extensive testing, reporting, contact tracing, and genome sequencing. The time-varying reproduction number can be estimated using various data sources, including case count data, contact tracing data, and genome sequence data. Stadler et al. [1] introduced a phylogenetic-based method using a birth–death model (BDM) process, which allows independent estimation of the basic reproduction number without making assumptions about the epidemiological properties of the pathogen. This work allowed, for the first time, independent estimation of ℛ_0_ from genomic sequence data alone. This approach inherits the limitations of the simplified assumptions of the BDM process, and the non-inclusion of epidemiological information can potentially limit the accuracy of ℛ_0_ estimates.

Studies have demonstrated that the estimation of the reproduction number can vary depending on the methods employed [2], and we anticipate that estimates will also vary based on the data source utilized. The sampling process can bias estimates of key epidemiological parameters like ℛ_0_ (basic reproduction number) and ℛ_*t*_ estimates from sequence data [3]. Therefore, identifying the data category that provides the most accurate estimates of the reproduction number under different scenarios is important. Recently, efforts have been intensified to improve the estimation of epidemiological parameters using a combination of genomics and epidemiological surveillance data [4–6]. In many settings, resources may be severely limited for simultaneous extensive epidemiological and genomic surveillance [7, 8]. In such situations, knowing which data sources will provide more accurate estimates may help in targeting scarce resources effectively.

Accurately predicting the time-varying reproduction number (ℛ_*t*_) is important for effective epidemic management. At a given time, an estimated ℛ_*t*_ near the critical value of 1 can misleadingly suggest either a decline or an increase in epidemic growth, depending on whether the true ℛ_*t*_ is slightly below or above this threshold. Precise estimation of ℛ_*t*_, when it is a proxy for the basic reproductive number, is also relevant for determining the necessary level of vaccination coverage to control an outbreak. For instance, if the estimated ℛ_*t*_ is 1.2 but the true value is 1.5, this represents a 25% error, but an ℛ_*t*_ of 1.2 translates to an estimated required vaccine coverage of 16.7%, compared to the required coverage of 33.3% if ℛ_*t*_ is 1.5. This corresponds to almost a 100% error in vaccine coverage prediction.

In this paper we address the question: Between genomic sequence data and case count data, which data source is likely to provide the best estimate of ℛ_*t*_, under a range of sampling conditions? To address this, we developed a simulation platform called ‘OOPidemic’ in R [9], which uses the SEIR modelling framework (susceptible-exposed-infectious-recovered) to simulate outbreaks and generate a genomic sequence for every infected individual in the outbreak, as well as a full linelist. Although other simulation packages have been developed, few are designed using the simple structured compartmental models commonly used in mathematical epidemiology, and therefore often lack the ability to easily simulate from specific epidemiological distributions of interest e.g. serial intervals and generation times. For example, FAVITES is an adaptable model based on transmission and contact networks [10] and e3SIM uses a more detailed agent-based approach [11]. Many methods simulate either the epidemiological or evolutionary dynamics first, and then layer in the other in a two-step process; this misses the important interactions between disease dynamics and pathogen evolution. An exception is the SEEDY package [12], which is no longer maintained on CRAN. We sought to develop a simple and fast simulation package, in R to facilitate easy onward analysis, using a basic evolutionary model but incorporating epidemiological quantities relevant to ℛ_*t*_ estimation and other transmission analyses.

This framework provides an opportunity to compare ℛ_*t*_ estimates from epidemiological and genomic sequence data where the ground truth is known. Although previous work has compared ℛ_*t*_ estimates from these data sources [13], as far as we know, they have not been compared to a ground truth under wide-ranging sampling scenarios.

## Materials and methods

Various methods have been proposed for estimating epidemiological parameters from sequence data [14] and epidemiological surveillance (case count) data. We chose the Birth-Death Skyline Model (BDSKY) from BEAST 2.7 as it is the most widely used in practice [15]. Post-processing of the data was done using the”bdskytools” and “beastio” package in R 4.3. Phylogenetic trees of the populations were visualized using DensiTree 2.7. Similarly, we chose EpiEstim [16], commonly used in practice for ℛ_*t*_ estimates for case count data. We aim to generate realistic sampling scenarios with known ℛ_*t*_, and compare commonly used tools.

We developed a simulation platform as an R package named ‘OOPidemic’ that uses the SEIR (Susceptible-Exposed-Infectious-Recovered) framework to simulate disease transmission with a fast object-oriented programming (OOP) approach. The simulator takes as input transmission rates, gamma distributed epidemiological parameters (recovery rate, incubation period, serial interval/generation time), the mutation rate, and population structure (size, initial infectives and strains, and optional group structure). As hosts are infected during the simulation, the package tracks which individuals are infected, by whom at what time, and when they become infectious and recover. It assigns each host a pathogen genomic sequence using a simple Jukes-Cantor evolutionary model. This setup generates a linelist for the simulated outbreak as a CSV (Comma-Separated Values) file and a FASTA file for the nucleotide sequences for all infected hosts. Generating these datasets in these formats makes it possible to easily load and process them in BDSKY and EpiEstim, respectively, in a straightforward manner. For each simulated outbreak, we used the entire linelist to obtain the true ℛ_*t*_ by calculating the average number of new infections generated by each host, infected on a specified day, throughout their infectious period. We further applied a smoothing function with a seven-day window to reduce the noise level in the true ℛ_*t*_ estimate.

A full description of the simulation model is available in the Supplementary Materials, and the ‘OOPidemic’ package is available on GitHub [17]. In Figure 1, we show a demonstrative simulated outbreak, with gamma-distributed incubation period, infectious period and serial interval, respectively, of mean 2, 5.5 and 3.5 days, and transmission rate 0.4 among all individuals. Further examples with different population sizes and more complex mixing structure are included in the Supplementary Materials. A)

**Fig 1.**
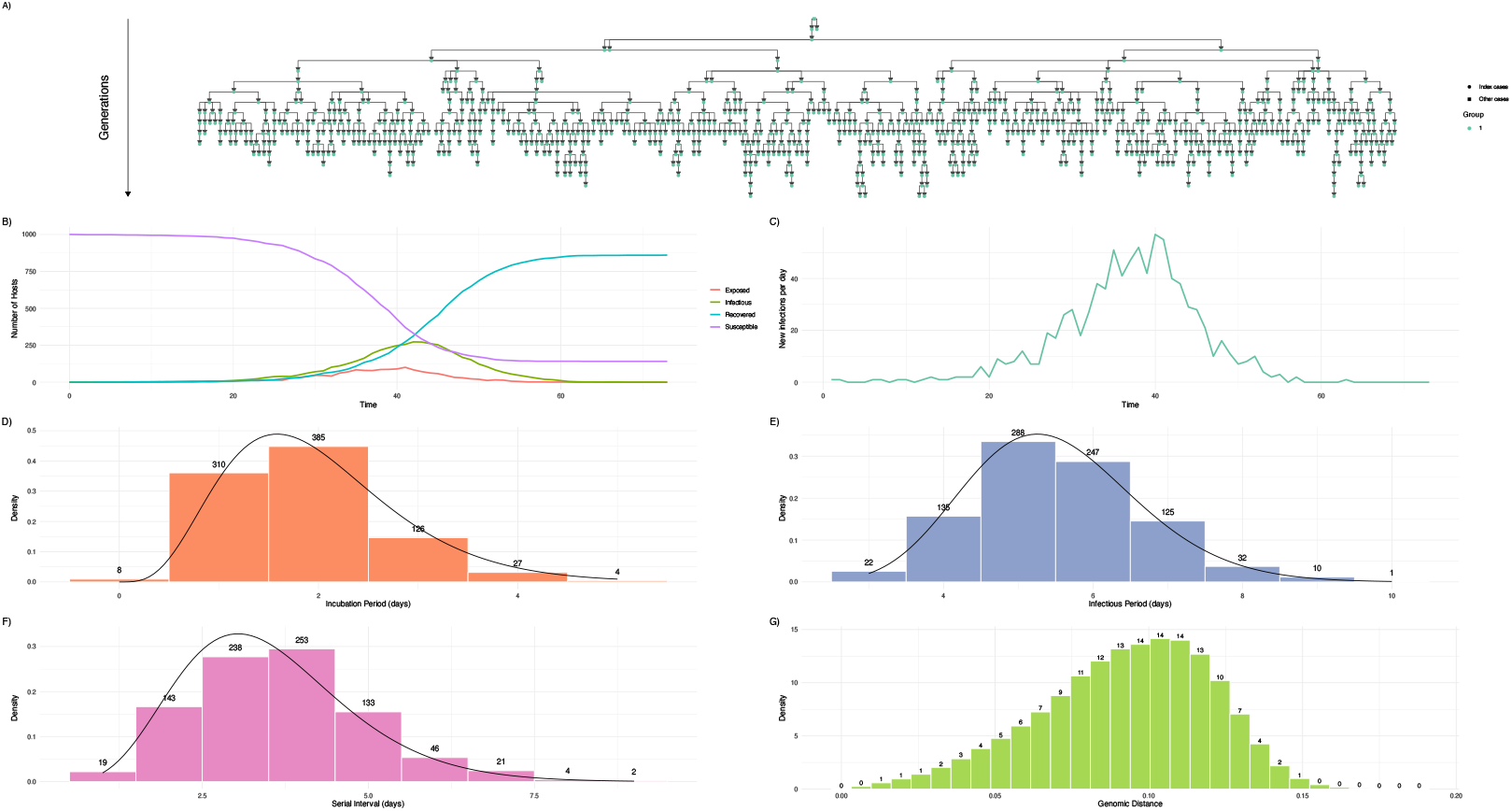
Demonstrative simulated outbreak of size 860 in a heterogeneously mixing population with 1000 hosts and 1 initial infector. Panels show the simulated (A) transmission tree, (B) compartment dynamics, (C) incidence curve, (D) distribution of incubation periods, (E) distribution of infectious periods, (F) distribution of serial intervals and (G) distribution of genomic distances between all possible pairs of infected hosts. In panels D, E, F, solid black lines show the gamma distributions simulated from.

We developed an automated pipeline in R that processes the linelist and generates the incidence and generation time distributions from the linelist, preparing them for ℛ_*t*_ estimation in EpiEstim. The EpiEstim package is designed to notify users when the data is insufficient at the start date of the estimation to reliably calculate ℛ_*t*_. We set up a process that determines an appropriate start date for ℛ_*t*_ estimates, ensuring that the data is sufficient for a reliable estimation.

Using the ‘OOPidemic’ simulation platform, we generated 20 outbreaks with different transmission rates, population size and infection seed, all of which are chosen within biologically meaningful bounds as shown in Table 1.

**Table 1.**
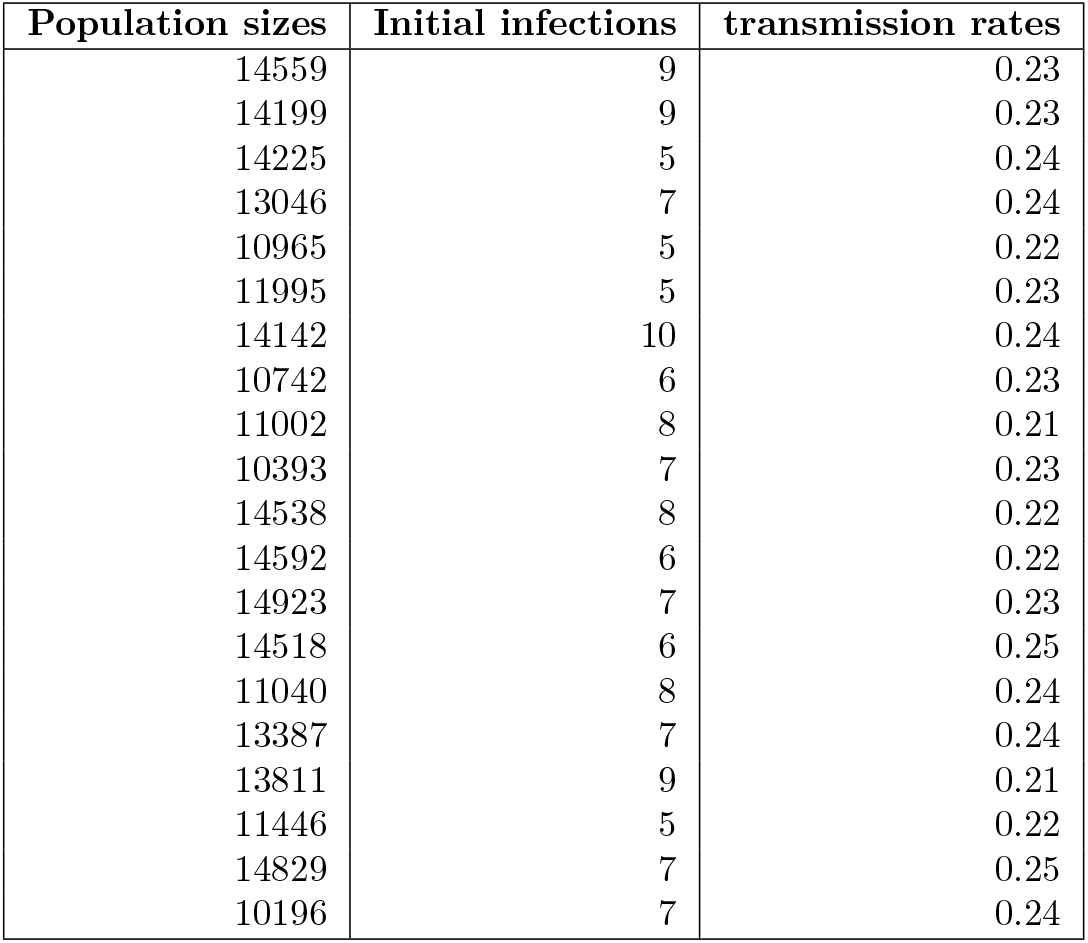
Showing values for population size, infections seed (initial infection), and transmission rates, for the simulated outbreaks. The duration of infection is gamma distributed with shape = 20 and rate = 4, translating in mean 5 days and variance 1.25 days.

The phylogenetic trees generated from the outbreaks are shown in Fig. 23. We created five scenarios to compare BDSKY and EpiEstim estimates. First, we considered a “complete sampling” scenario called Scenario A. For the EpiEstim estimate, we obtained the daily case count and the generation times from the linelist and used them as input to the EpiEstim package in R to generate ℛ_*t*_ estimates. On the other hand, due to the computational costs of using sequences for all infected hosts, we downsampled gradually and compared ℛ_*t*_ estimates until we obtained an optimal number of sequences, above which adding more sequences did not significantly change the BEAST estimates (but below which the estimates are different). This process allowed us to identify a “complete” sequence count per day. We repeated this for multiple test examples to determine an agreed-upon proportion for Scenario A, which we used as the benchmark for downsampling in other scenarios for the BDSKY estimates.

Second (Scenario B), for EpiEstim, we considered uniform downsampling of 70% of the entire linelist; for each day, we sampled a number of hosts following a negative binomial distribution. To achieve this, we counted the number of cases per day, denoted as *n*, and multiplied it by the sampled fraction *f* (e.g., 70%). We then calculated the average number of cases per day after downsampling, denoted as *µ*_*f*_. To correctly estimate the dispersion parameter *k* of the negative binomial distribution, we first computed the mean and variance of the original daily case counts, denoted as *µ* and *σ*^2^, respectively.

The dispersion parameter *k* was then estimated using:

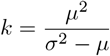

where *k* controls the level of overdispersion in the data. Using *µ*_*f*_ as the mean and *k* as the dispersion parameter, we generated the downsampled case counts by sampling from a negative binomial distribution:

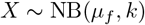

This ensures that the downsampled case counts reflect both the mean reduction due to subsampling and the overdispersion in the data.

We followed a similar process for the BDSKY estimate, but only sampled some of the sequences. We selected 400 sequences as a feasible number to run in BEAST within a reasonable timeframe. In Scenario A (equivalent to the EpiEstim estimate Scenario A), all time points are sampled according to this rate. If a time point has fewer individuals than required, one individual is randomly selected to ensure representation from every time point. For the BDSKY estimates, all other down-sampled scenarios are done with reference to Scenario A and not the raw numbers due to computational limitations. Unlike in Scenario A, not all time points are sampled in the down-sampled scenarios. Table 2 shows the number of sequences used in each population and scenario. Third (**Scenario C**), we repeated the same process as Scenario B for the EpiEstim estimate, but this time sampling, on the average, only 10% per day. We were interested in observing how reducing the uniform sampling proportion would impact the estimates from both approaches. Fourth (**Scenario D**), we considered scenarios where the sampling proportion changes as the epidemic progresses by introducing a switch point. We considered where the sampling proportion increases from 10% to 70% as the outbreak approaches its peak. This is akin to situations where testing and sequencing capacities are ramped up as the epidemic grows. Fifth (**Scenario E**), we considered a sampling scenario where the sampled proportion decreases from 70% to 10%, with the change following a sigmoidal function over a few days.

**Table 2.** Number of sequences that were used in each population/scenario.

| Pop\Scenario | Raw | Scenario A | Scenario B | Scenario C | Scenario D | Scenario E |
| --- | --- | --- | --- | --- | --- | --- |
| 001 | 5844 | 454 | 286 | 41 | 116 | 219 |
| 002 | 5691 | 459 | 279 | 40 | 139 | 196 |
| 003 | 6065 | 485 | 297 | 42 | 128 | 221 |
| 004 | 5666 | 448 | 278 | 40 | 104 | 212 |
| 007 | 6576 | 432 | 276 | 39 | 128 | 191 |
| 008 | 4356 | 427 | 274 | 39 | 120 | 198 |
| 009 | 3235 | 446 | 272 | 39 | 105 | 216 |
| 010 | 3994 | 428 | 280 | 48 | 112 | 282 |
| 012 | 4597 | 470 | 290 | 41 | 130 | 191 |
| 013 | 6105 | 473 | 299 | 43 | 134 | 202 |
| 014 | 7030 | 492 | 295 | 42 | 128 | 212 |
| 015 | 4637 | 469 | 292 | 42 | 97 | 229 |
| 016 | 5367 | 423 | 263 | 38 | 111 | 195 |
| 017 | 4198 | 478 | 294 | 42 | 147 | 176 |
| 018 | 4278 | 441 | 270 | 39 | 141 | 172 |
| 019 | 7385 | 416 | 258 | 37 | 131 | 167 |
| 020 | 4336 | 426 | 273 | 39 | 144 | 181 |

To compare the performance of these approaches in the various scenarios listed above, we employed the Dynamic Time Warping (DTW) method [18], which measures similarity between time series, even if the series are time-shifted and/or of different lengths. DTW uses a warping path through a grid that minimizes the cumulative distance between points in the two time series, using Euclidean distance to compute the distance between points. This path represents the optimal alignment between the two time series. Due to its complexity, the DTW approach may present some difficulties for interpretation. To build intuition for its interpretation, in Fig 2, we show multiple time series and the application of DTW to compare them. Time series one is the reference series, and the other series are compared to it. The level of alignment of the black lines with the dotted red lines, in the second column of Fig 2, shows how similar each series is to the reference series.

**Fig 2.**
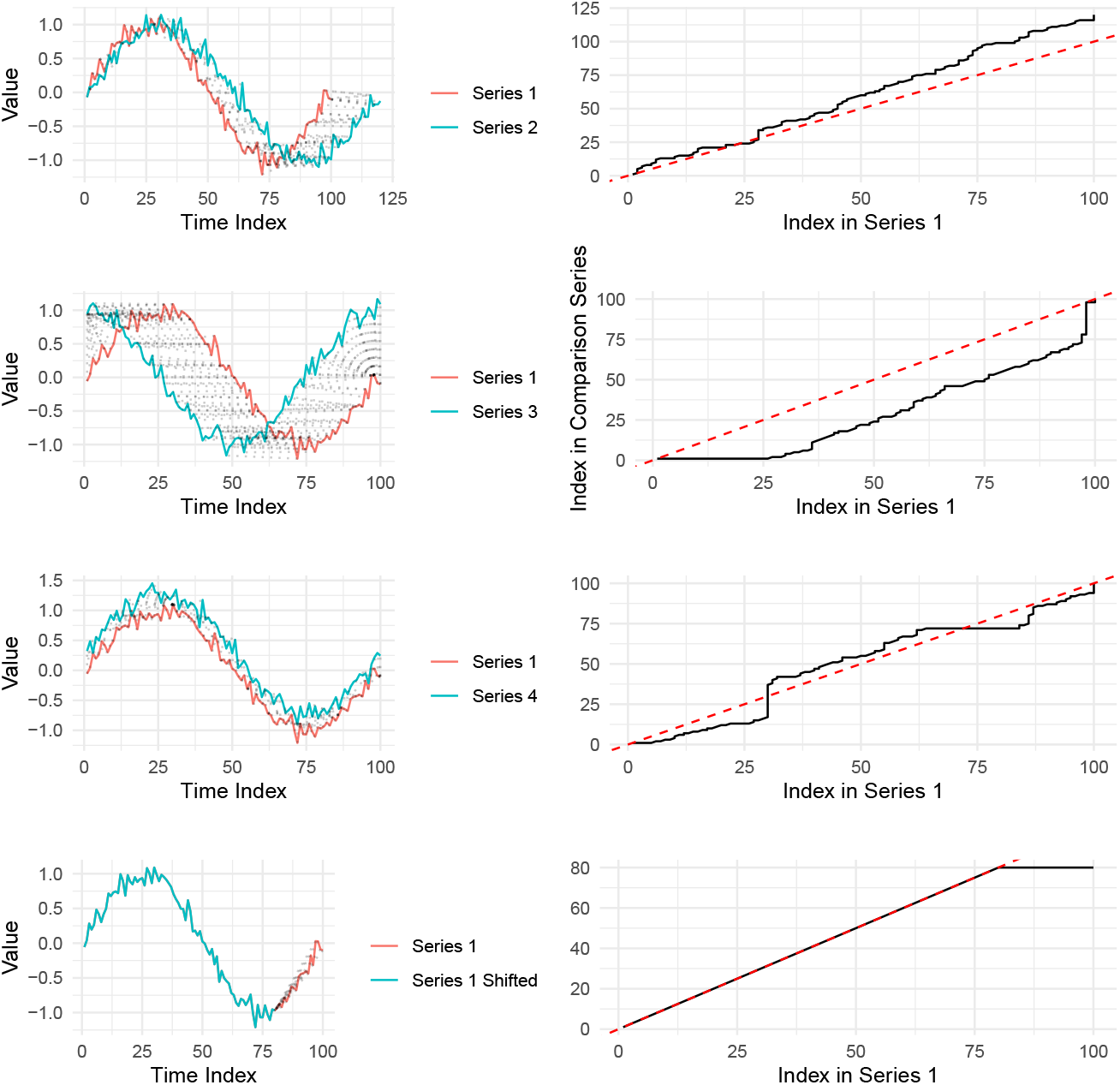
Sample time series and application of DTW for comparison. The first column shows four series; series one is the reference series, which is compared to series two, three, four and shifted time series one. The gray dotted lines are the warping paths. The second column shows the application of DTW for comparison. Diagonal dotted red lines indicate perfect alignment when two time series are exactly the same. The black lines show the alignment between each of the time series and series one (the reference time series).

We computed the Root Mean Square Error (RMSE) to quantify the deviation of the EpiEstim and BDSKY estimates from the reference ℛ_*t*_ estimate, providing a quantitative measure of accuracy in each scenario (Fig. 8). Similarly we calculated the median RMSE for each scenarios to show how the accuracy of each estimation approaches changes for various scenarios (Fig. 8, bottom left panel). In the Results and Discussion sections, we used EpiEstim estimates and casecount estimates interchangeably, as well as BDSKY estimates and sequence estimates.

## Results

We analyzed the linelist and the sequence data for each simulated outbreak using EpiEstim and BDSKY, respectively. Figure (3) shows the results for Scenario A; we found that in this scenario, the EpiEstim estimate is consistently better aligned with the true ℛ_*t*_ value. The sequence-based approach underestimates ℛ_*t*_ in the first part of the outbreak and overestimates it in the latter part. On the other hand, the case count estimates (from EpiEstim) are consistently well aligned with the true ℛ_*t*_. The level of alignment between the estimates and line denoting a perfect match between time series (labelled ‘Perfect align’ in Figure 3) indicates the level of accuracy of the estimates. In Figures 3–7, each row represents an analysis of an outbreak generated by different transmission and recovery rates. Note that in scenarios other than the optimum case, the ℛ_*t*_ estimate from the sampled sequences may not extend to the final time point of the simulation. This is because, with uniform sampling, not all time points could be selected. In other words, the ℛ_*t*_ estimate only extends to the last time point that was randomly sampled. The RMSE estimates in Fig. 8 show a consistent pattern, with the BDSKY estimates having a higher RMSE for all outbreaks included in the analysis. This result agrees with the DTW alignment for all outbreaks.

**Fig 3.**
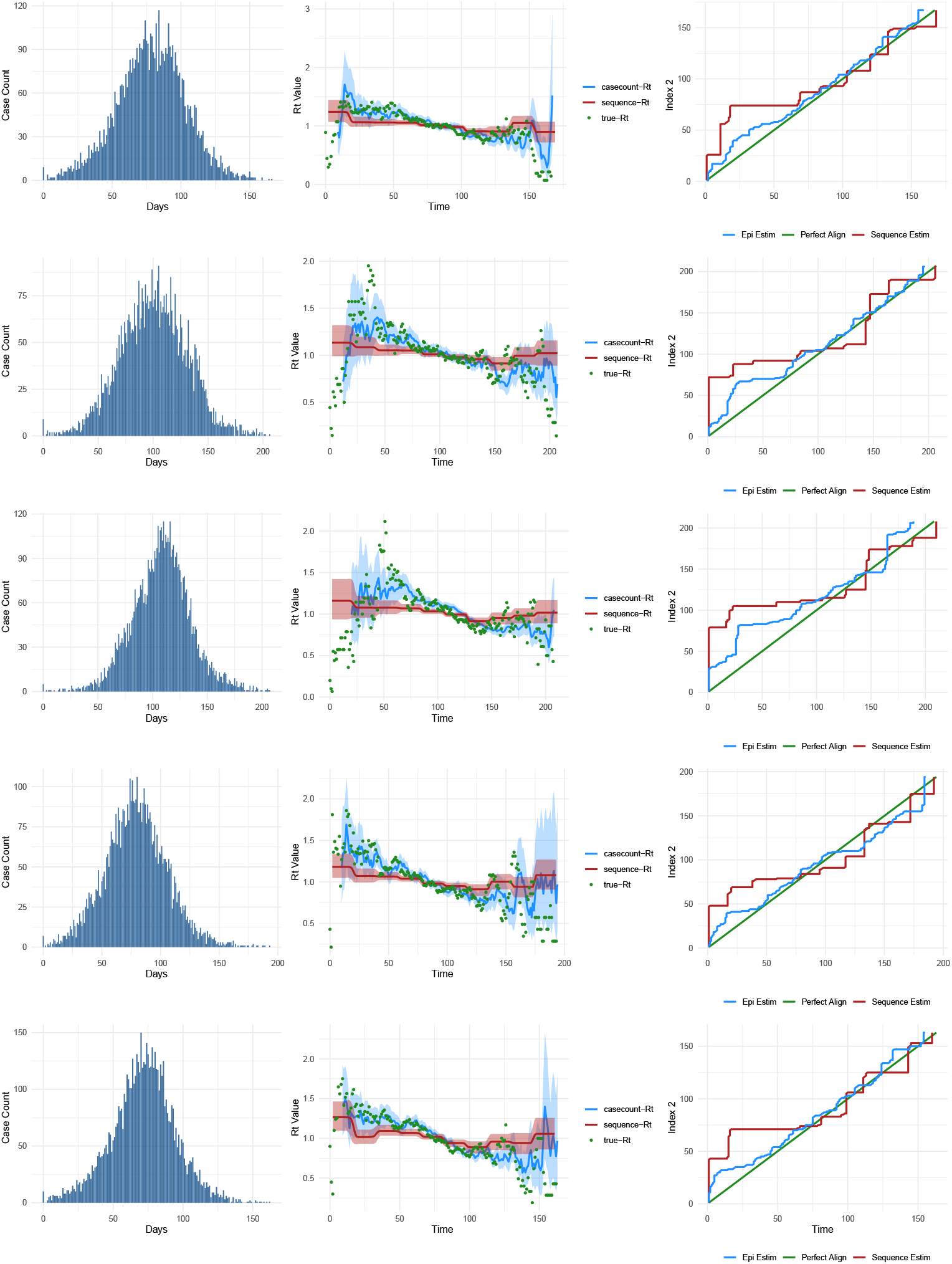
Outbreaks and comparison of accuracy of ℛ_*t*_ estimates from case count data and sequence data for Scenario A. The first column (bar plots) shows the incidence of cases per day over the course of the outbreak. The second column shows the estimates from the two data sources compared to the true ℛ_*t*_ estimate. The third column shows the alignment of each estimate with the true estimate. The green line represents the perfect alignment. Each row represents a simulated outbreak.

We applied the DTW methods to compare estimates for Scenario B, keeping an average of 70% of the Scenario A per day (Fig 4). The case count estimate still yields better estimates than the sequence-based method. Both estimates are worse in Scenario B than in Scenario A (see the middle and last columns in Fig 4). We observed a similar pattern in the RMSE results, with minimal difference in the case count estimate between Scenarios A and B compared to the sequence estimate with a more pronounced difference (Fig. 8). In the 10% downsampled scenario (Scenario C; Fig 5) the estimates are yet worse, as expected, and are both not as accurate as they are in Scenarios A and B. Both estimates tend to follow the same pattern – for outbreaks where one aligns well with the true ℛ_*t*_, the other also tends to have good alignment and vice versa. This is evident both by visual inspection of the estimates and the DTW alignments in the third column of Fig 5. We computed the RMSE in this scenario, and it has a higher value for both the case count and sequence estimates compared to Scenarios A and B (Fig. 8). Moreover, it is evident in Fig. 8 that the case count estimate is closer to the truth than the sequence estimate in Scenario C. We also notice a consistently higher overestimation for the sequence-based estimate earlier in the data, which might be due to the sequence estimate underestimating diversity because of the low number of sequences and interpreting delayed coalescent events as high transmission, or it might be due to overfitting noise in the data.

**Fig 4.**
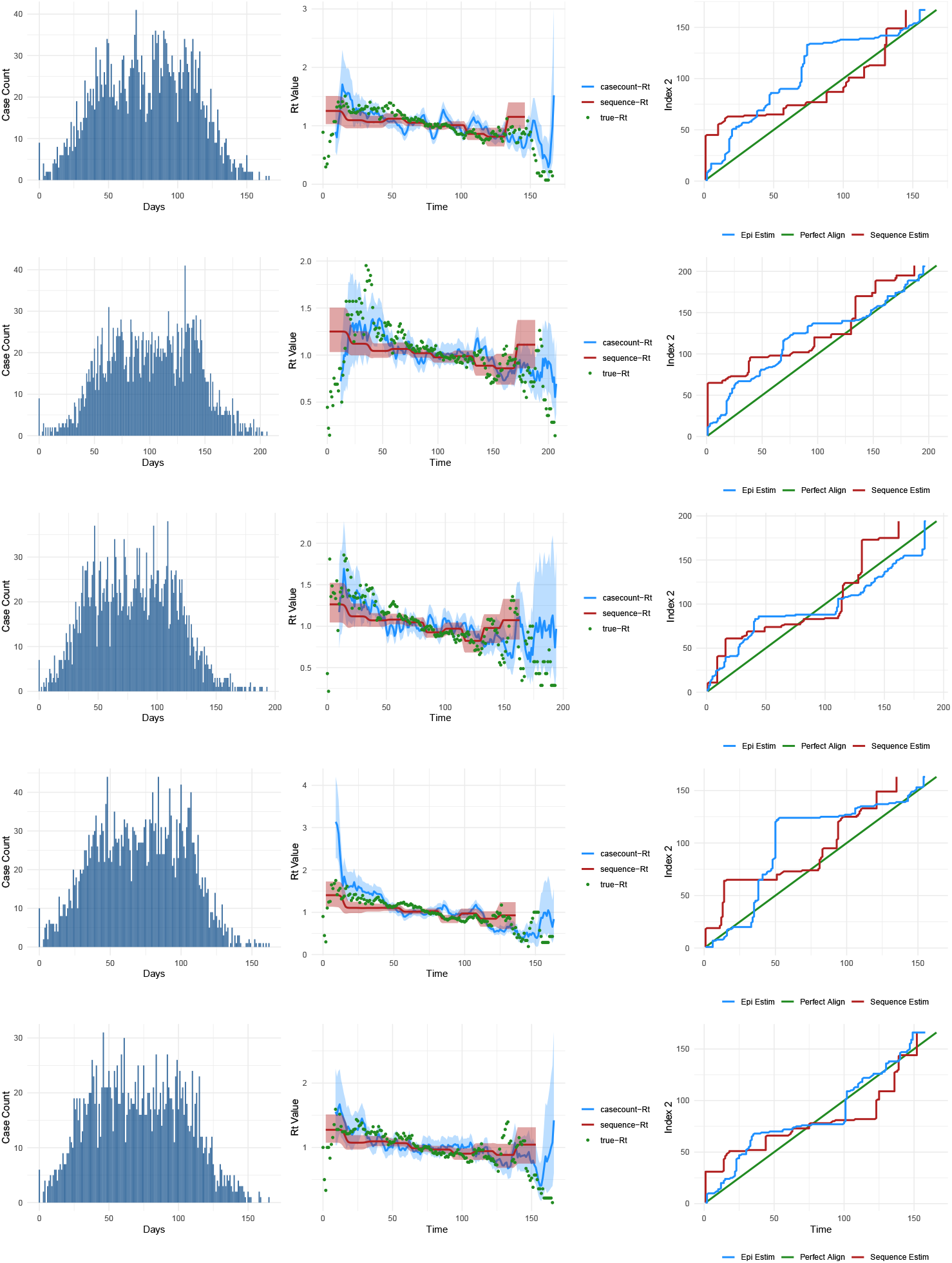
Outbreaks and comparison of the accuracy of ℛ_*t*_ estimates from case count data and sequence data for scenario B (70% sampled)). The first column (bar plots) shows the incidence of cases per day over the course of the outbreak. The second column shows the actual estimates from the two data sources compared to the true ℛ_*t*_ estimate. The third column shows the alignment of each estimate with the true estimate. The green line represents perfect alignment.

**Fig 5.**
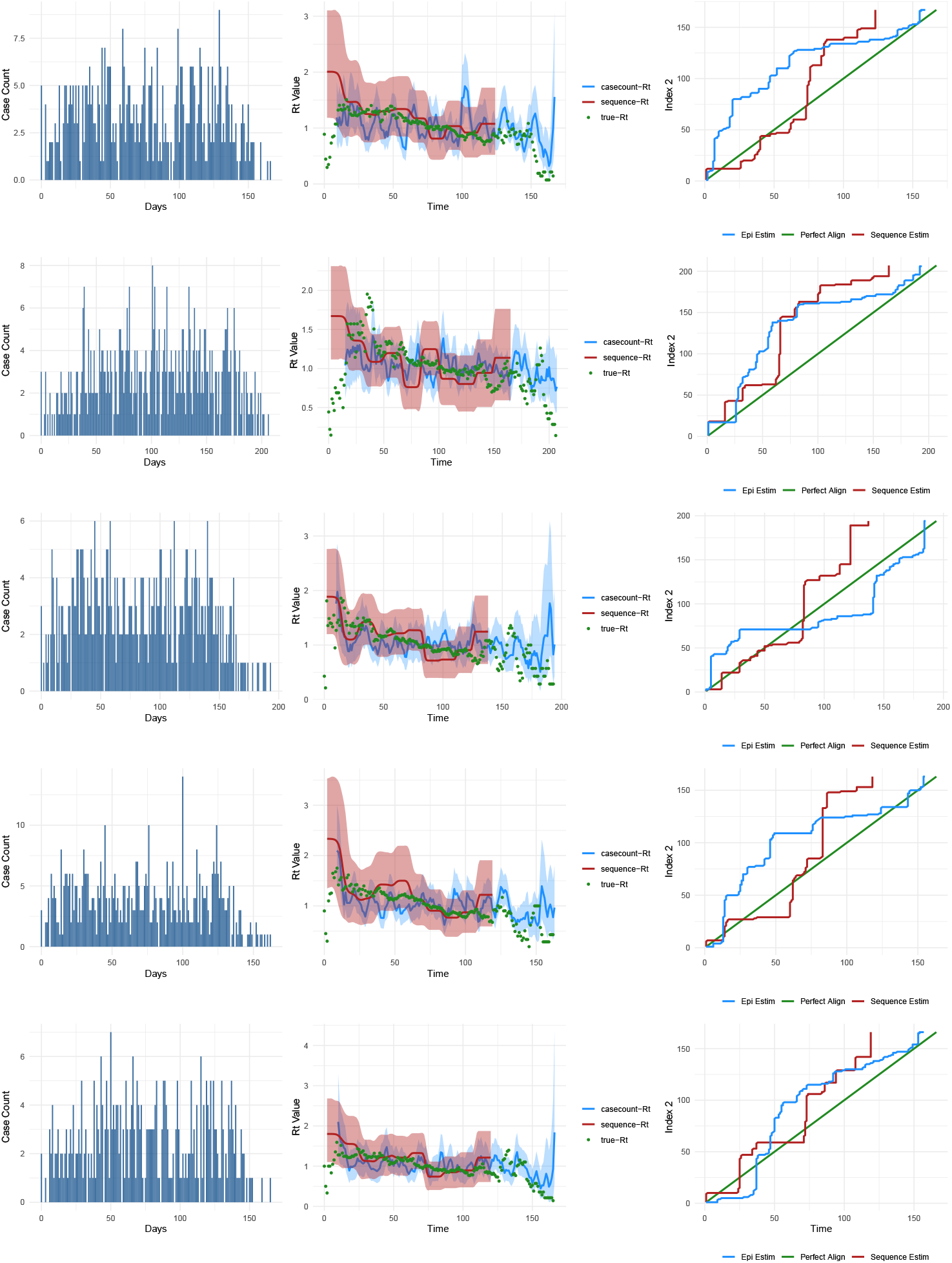
Outbreaks and comparison of the accuracy of ℛ_*t*_ estimates from case count data and sequence data for Scenario C (10% sampling scenario). The first column (bar plots) shows the daily incidence of cases over the course of the outbreak. The second column shows the estimates from the two data sources compared to the true ℛ_*t*_ estimate. The third column illustrates the alignment of each estimate with the true estimate. The green line represents perfect alignment.

For the previous sampling scenarios (Scenarios B and C), we have looked at uniform sampling throughout each of the outbreaks. We now consider a switch during the course of an outbreak, where the sampling proportion changes from low (10%) to high (70%) before the peak of the outbreak (Scenarios D). The sequence estimates, although not perfectly aligned, have a clearly far better alignment than the case count estimates (Fig. 6). The RMSE also shows a similar pattern, with consistently higher values for the case count estimates and lower RMSE for the sequence estimates. This holds true for all outbreaks simulated in this analysis (Fig. 8). Although the case count estimates improve after 50 days when more data become available, and become comparable to the sequence-based estimates, the sequence estimates perform better overall in this scenario.

**Fig 6.**
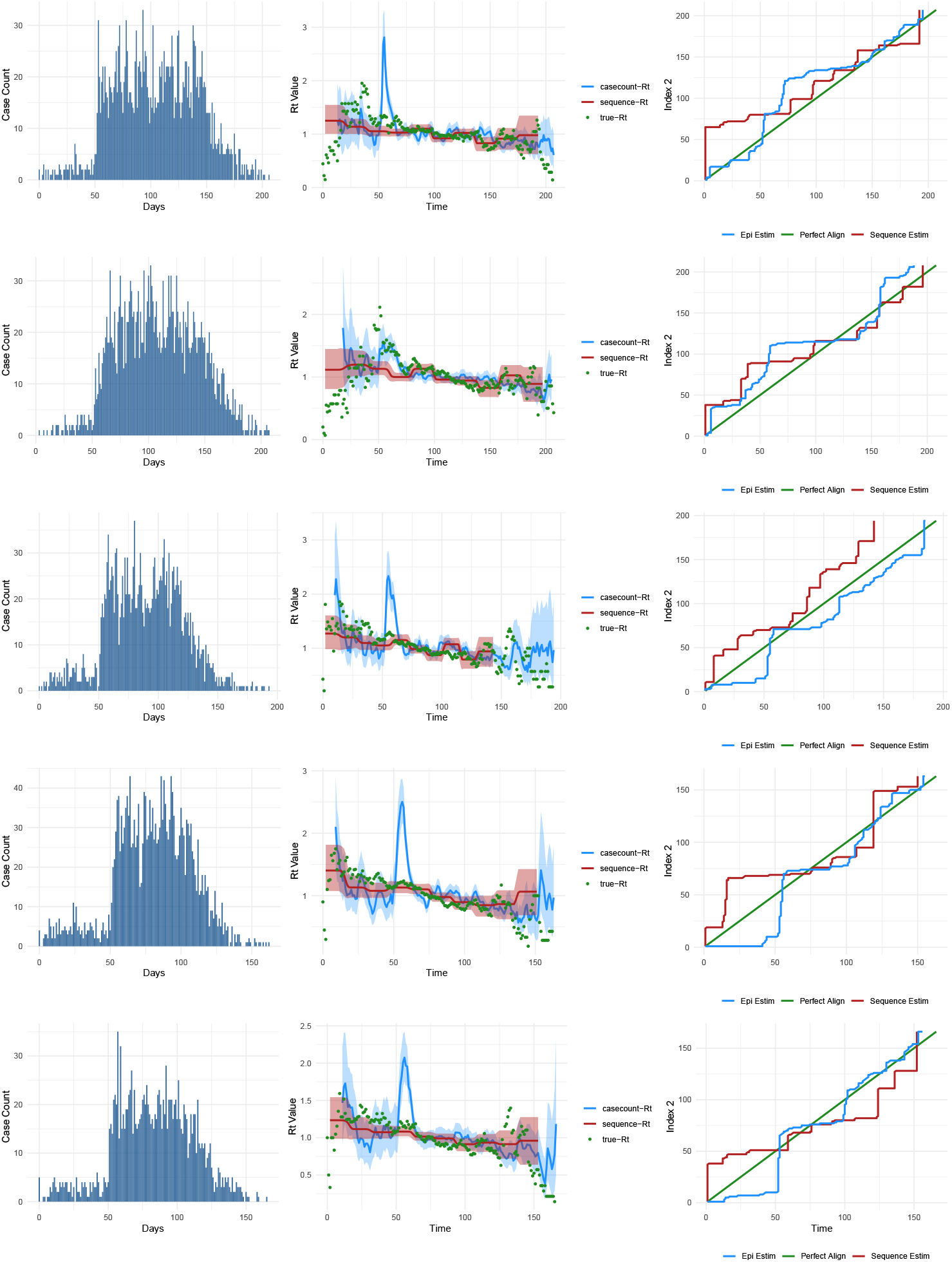
Outbreaks and comparison of the accuracy of ℛ_*t*_ estimates from case count data and sequence data for Scenario D (10% to 70% sampling scenario). The first column (bar plots) depicts the daily incidence of cases throughout the outbreak. The second column presents the estimates from the two data sources in comparison to the true ℛ_*t*_ estimate. The third column demonstrates the alignment of each estimate with the true estimate. The green line represents perfect alignment.

We created a scenario with a switch from high (70%) to low (10%) sampling proportion (Scenario E). This is plausible during outbreaks in situations where testing and tracing capacities suddenly break down due to limited resources, a sudden increase in the number of cases beyond existing capacities, a change in testing policies, or a combination of these factors. In Fig 7, the case count estimates are not aligned in all of the simulated outbreaks. By visual inspection of the sequence comparison plot, the case count estimates are mostly inaccurate, especially near periods when the data are significantly reduced, and for most time points across the simulated outbreaks, they deviate from the true estimate. However, the performance of the sequence estimates, although more consistent over time, is not better than that of the case count estimates overall, as they deviate from the true ℛ_*t*_ at the tail. Fig 7 shows Scenario E – the downsampled simulated outbreaks, the estimates from the two data sources, and the DTW alignments for multiple outbreaks. Case count ℛ_*t*_ estimates consistently show better alignment with the true estimates compared to the sequence data estimates for all the included outbreaks. The estimates from the sequence data did not align very well with the true estimates when compared to the case count estimates, but the alignment is not visibly worse than the estimates from the 10% sampling scenario. In fact, RMSE shows that sequence estimates is slightly more accurate in Scenario E compared to Scenario C (Fig. 8). Apart from Scenario C, the performance of sequence estimates is generally comparable. On the other hand, Scenarios A and B are comparable for the case count estimate, while Scenarios C and D are also fairly comparable. We also added tree visualizations from the BEAST estimates for the 20 outbreaks, as well as for Scenarios A to E (see Fig. 23 in the supplementary information). Additionally, larger figures containing more scenarios and outbreaks, depicting the outbreak trajectory, ribbon plots for estimate comparisons, and the DTW alignment, are presented in Figs 18–21 of the supplementary information.

**Fig 7.**
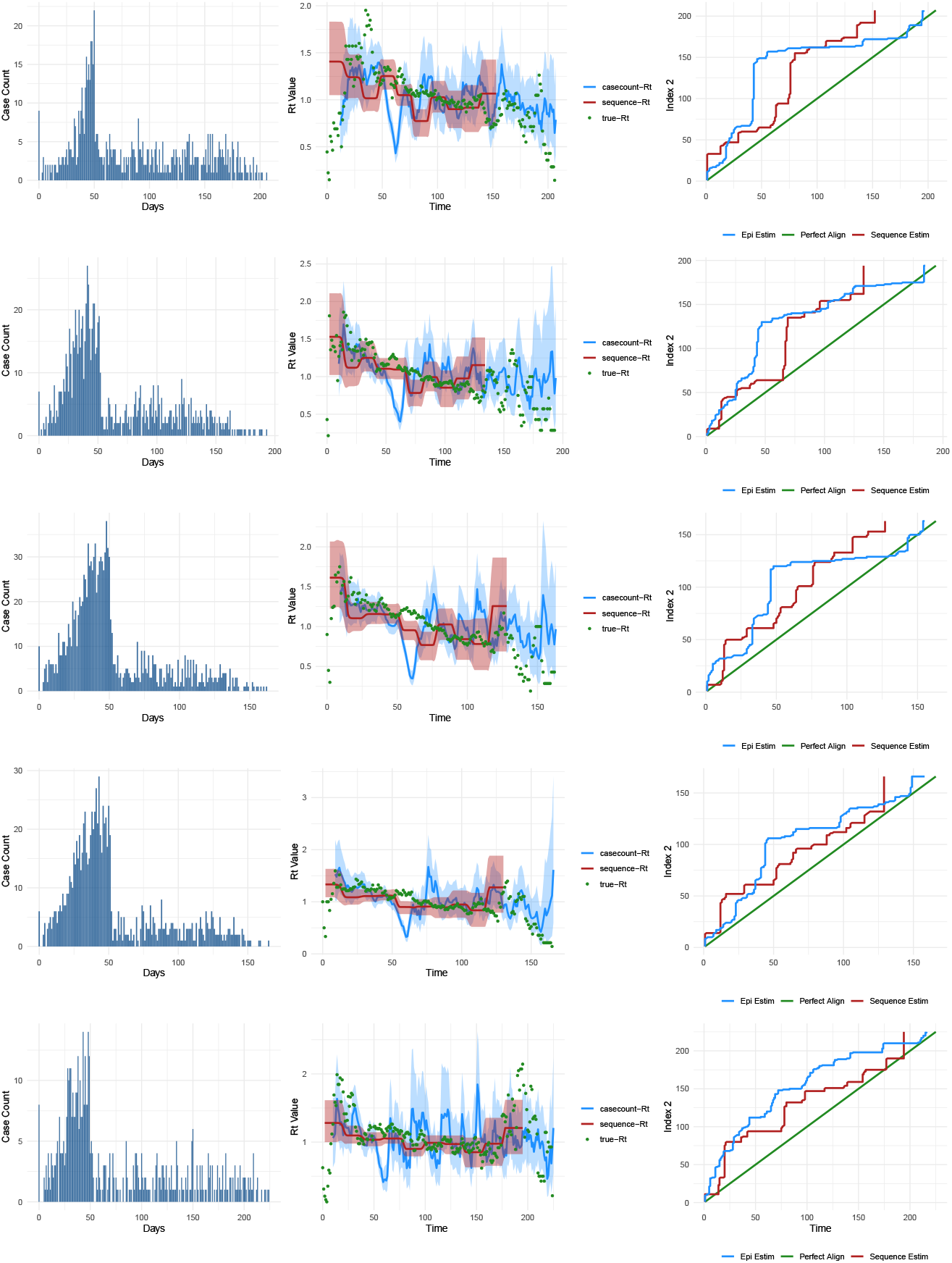
Outbreaks and comparison of the accuracy of ℛ_*t*_ estimates from case count data and sequence data for Scenario E (shift from 70% to 10% sampling scenario). The first column (bar plots) illustrates the daily incidence of cases over the course of the outbreak. The second column displays the estimates from both data sources compared to the true ℛ_*t*_ estimate. The third column shows the alignment of each estimate with the true estimate. The green line indicates perfect alignment.

**Fig 8.**
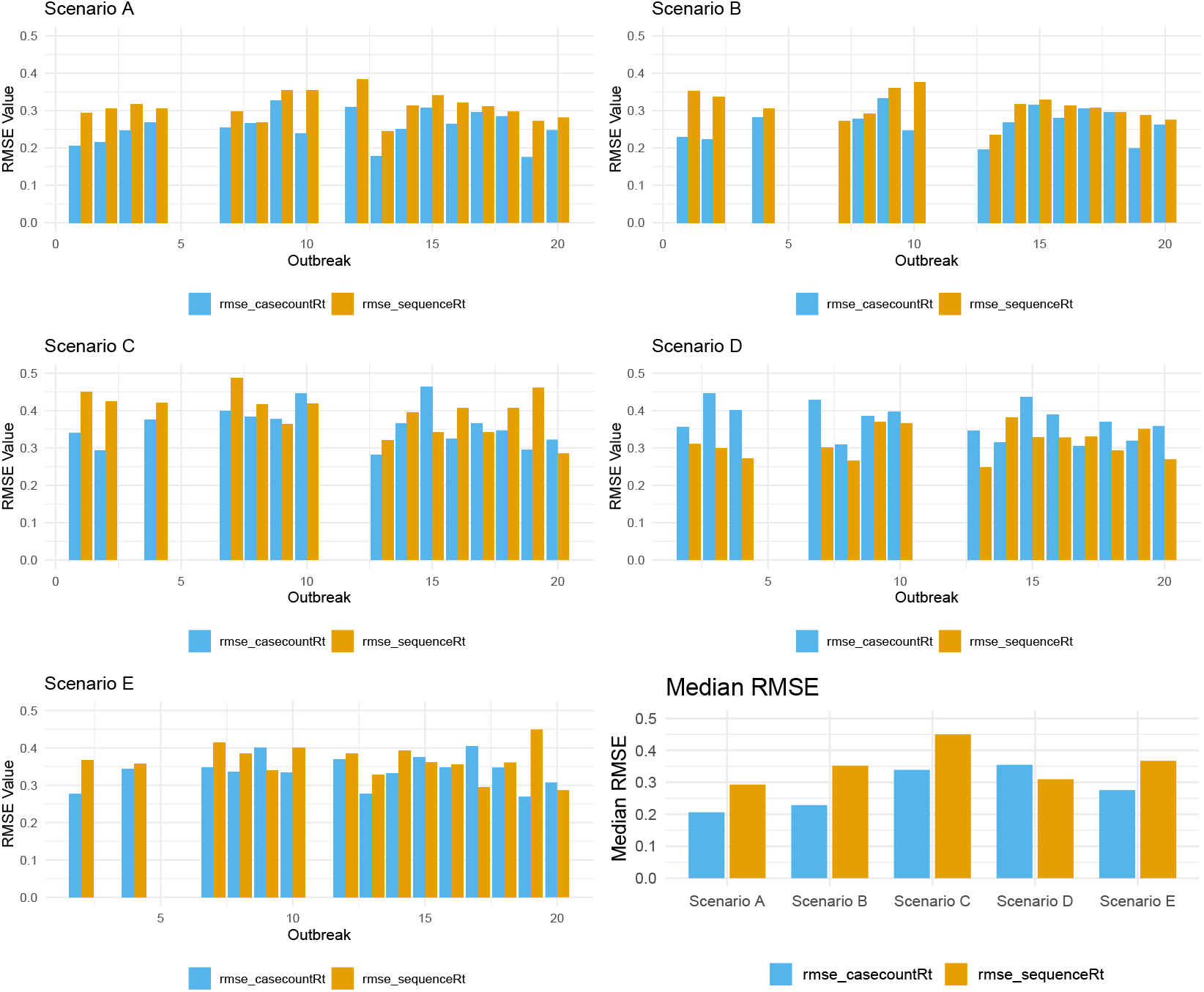
Bar plots showing the Root Mean Square Error (RMSE) for EpiEstim (case count) estimates and BDSKY (sequence) estimates compared to the true ℛ_*t*_ for Scenarios A, C–E. The upper left plot shows the RMSE grouped by outbreak number for Scenario A. The upper right plot shows the RMSE for Scenario C. The lower left plot shows the RMSE for Scenario D, and the lower right plot shows the RMSE for Scenario E. For each group, blue bars represent the RMSE for the EpiEstim estimates, and orange bars represent the RMSE for the BDSKY estimates. The bottom right panel shows the median Root Mean Square Error (RMSE) for Scenarios A to E. Each pair represents a scenario, color-coded by the case count (blue) and sequence (orange) estimates

## Discussion

As expected, the accuracy of ℛ_*t*_ estimates from sequence and case count data depends not only on the source of the estimates but also on the quality of the data. Moreover, we found that, as expected, the accuracy of case count estimates increases as more data is included. However, for sequence-based estimates, increasing the number of sequences does not always improve accuracy. In fact, our results show a scenario where a lower number of sequences yields more accurate ℛ_*t*_ estimates than a higher number, specifically with Scenario D being more accurate than Scenario B, despite having fewer sequences. This may, in part, be explained by a higher concentration of sampling in the later stages of the outbreak.

For both estimation methods, having a very low number of sampled cases markedly and negatively impacted the ℛ_*t*_ estimates. This finding partially aligns with previous studies on estimating population genomic parameters from sequence data, which suggest that increasing sample size generally improves accuracy [19, 20]. However, the benefits of increasing sample size may have trade-offs and do not scale linearly, as sampling biases can lead to situations where fewer sequences result in more accurate estimates.

The accuracy of case count estimates is high if the sampling is uniform and the downsampled data has a similar shape to the original outbreak data. However, the performance is unreliable in situations where insufficient testing may suggest that the epidemic is declining when it is actually growing, similar to the Omicron wave, where factors like changing testing strategies, reporting behaviors, and changes in disease severity may bias downward the reported cases [21]. This is a documented limitation of the EpiEstim method [16]. On the other hand, the performance of sequence estimates is fairly consistent save for the vary low sampling scenario, regardless of whether the testing bias affects the shape of the observed outbreak data. One insight from the phylogenetic trees is that in lower sampling regimes, where the number of sequences is smaller, the trees show less uncertainty (See Fig 23 in the Supplementary material), which may explain the better performance in estimation.

Generally, sequence data is obtained from ascertained cases, making it difficult to disentangle the qualities of the two data sources in practice. Studies like this can inform decisions on efficiently deploying resources, especially in settings with limited resources. Choices need to be made between focusing on wide testing and contact tracing to directly estimate ℛ_*t*_ or concentrating on sequencing the ascertained cases. Our findings suggest that when testing is unreliable, efforts should focus on sequencing the ascertained cases or adjusting testing strategies to improve the quality of surveillance data, yielding accurate estimates with case count-based methods like EpiEstim. The caveat is that sequencing provides a wide range of important information about the pathogen beyond ℛ_*t*_ estimates, such as inferring transmission chains [22] and selection coefficients. We did not extend our findings beyond the accuracy of ℛ_*t*_ estimates in different scenarios. These results should be interpreted in that context.

We did not use any real-world data in this study mainly because it is very challenging to have full information about all infected cases in the field, so using real outbreak data presents some unique challenges. There have been surveillance data that can be used, similar to [13], but full knowledge of exactly who infected whom and when necessitates the use of synthetic data. The study has several limitations. We did not compare a wide range of methods for estimating ℛ_*t*_ from either case count or sequence data. Very recently, efforts have been made to integrate case counts and sequence data into a single methodological framework, known as EpiFusion, to leverage both data sources. The results are promising and, similar to our study, indicate that each data source may have advantages depending on the context [23].

Moreover, given the clear distinction in accuracy across different scenarios, simple weighted averaging methods could provide a straightforward approach for combining estimates from both data sources while adjusting for biases where appropriate. The primary focus of this study was to evaluate commonly used methods and establish scenarios in which one approach proves more accurate than the other.

## Conclusion

We have shown that independent estimates of the time-varying reproduction number from sequence and case count data vary in accuracy depending on the data quality. When data is sparse and unreliable, sequence estimates, although not as accurate as case count data when data quality is optimal, offer better accuracy in such situations. We call for more studies leveraging these data sources to develop methods that will be robust to sampling biases and poor quality data, and still provide high accuracy. Increasing sequence sample size can be counterproductive due to financial and computational costs and may offer less accuracy beyond a certain threshold. More efforts should be put into finding optimal sequences in specific contexts to reduce costs and optimize accuracy.

## Data Availability

All data produced in the present work are contained in the manuscript

## Acknowledgments

We would like to thank Christopher Douglas for his work on the ‘OOPidemic’ simulation package.

## Funding

CC acknowledges support from the Canada 150 Research Chair, the Natural Sciences and Engineering Research Council of Canada (NSERC) Discovery Grant (RGPIN-2019-06624), and the Canadian Network for Modelling Infectious Disease (CANMOD). NSM also received support from the CANMOD. JS received support from the Natural Sciences and Engineering Research Council of Canada (NSERC) Discovery Grant (RGPIN-2023-4509). The funding bodies had no role in the design of the study, data collection and analysis, decision to publish, or preparation of the manuscript.

## Supplementary materials

### Simulation model

In this section, we detail the simulation model and approach used in the ‘OOPidemic’ outbreak simulator.

#### Epidemiological model

The base ‘OOPidemic’ model generates an SEIR outbreak in a closed, homogeneously mixing population. Susceptible (S) individuals may become infected via contact with infectious (I) individuals, at which point they become exposed (E). At the end of their incubation period, exposed individuals become infectious (I) and eventually removed (R). Removed individuals are fully protected with no waning immunity. The lengths of time spent in the E and I compartments are by default gamma distributed, with shape and rate parameters input by the user. It is simple to reduce the model to SIR by setting the length of the exposed periods to 0. The simulator requires the user to define a serial interval distribution, or alternately a generation time or transmission interval distribution (all gamma distributed), as summarized in Fig. 9. These distributions are used to find an infector for each infectee, as will be described below. We make the simplifying assumption that onset of symptoms and onset of infectiousness are simultaneous, at entry to the infectious compartment I. This model is not designed for settings in which transmission intervals are expected to often be negative: in fact, the simulator will never draw negative transmission intervals. If the chosen parameters produce negative transmission intervals more than 50% of the time, a warning will be returned to the user, and the distribution of the transmission interval will be truncated at 0.

**Fig 9.**
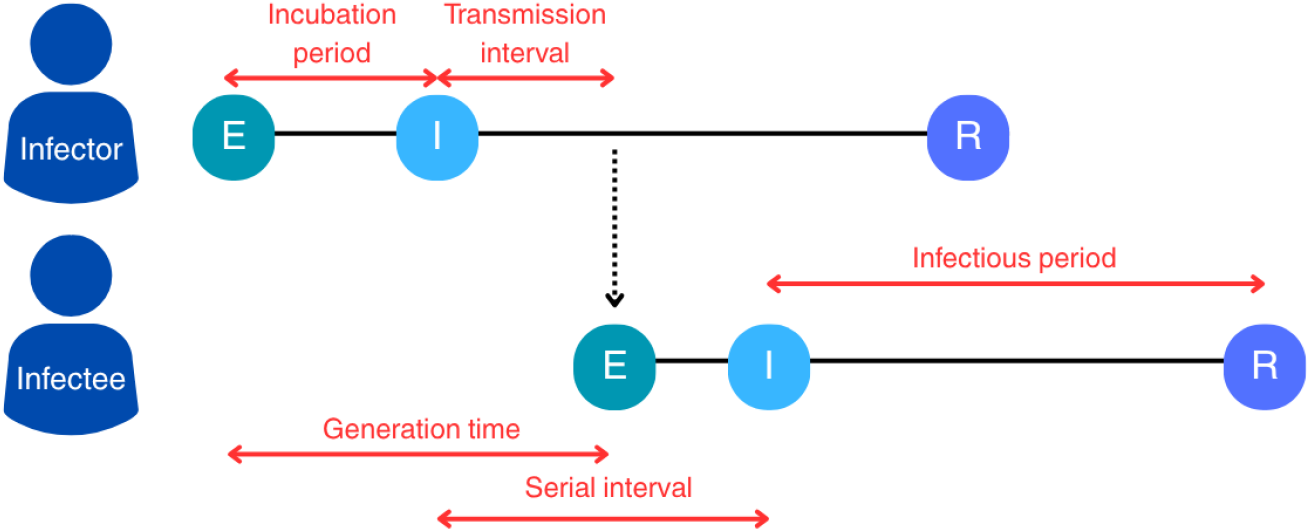
Transmission model schematic. For any (infector, infectee) pair, the transmission interval defines the length of time from onset of infectiousness in the infector to onset of infectiousness in the infectee. The generation time describes the length of time from exposure of infector to the exposure of infectee, and the serial interval describes the length of time from onset of infectiousness in infector to onset of infectiousness in infectee. All time periods depicted are assumed to be gamma distributed in the ‘OOPidemic’ model.

To initialize a simulation, users must build a ‘Group’ object - defining the initial numbers of susceptible and infected individuals, the infection rate and the above epidemiological distributions. The model is individual-based, that is, building the Group object automatically generates one ‘Host’ object per individual. Each Host object retains a full trace of its epidemiological trajectory, including compartment transitions, sampling times, infector and infectees, and mutation history. This allows detailed downstream reconstruction of the transmission tree and evolution across the outbreak.

Simulations proceed in discrete time steps until no exposed or infectious hosts remain. The number of new infections at each time step is drawn from a binomial distribution, based on the calculated force of infection

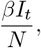

given transmission rate *β, I*_*t*_ infectious individuals at time *t*, and total population size *N*. Since all individuals receive the same force of infection in the homogeneously-mixing model, the specific individuals to be infected are selected uniformly at random. Upon infection of a new host, this host will have their future times of infectiousness onset and removal simulated by drawing from the relevant gamma distributions. The model samples a serial, generation, or transmission interval (whichever was defined by the user) for the new host by drawing from the respective gamma distribution. Lastly, a future time for sampling is generated for each host, by default at a uniform random time during their infectious period. The package also includes the option (i) to set regular intervals at which all currently-infectious hosts are sampled, or (ii) to sample all infectious hosts at fixed time intervals from infection until recovery.

The simulator additionally tracks the infector of each infected host. In the case where a serial interval distribution was input by the user, all feasible infectors (those currently infectious) will have their own sampled serial interval, as described above. The model selects whichever feasible infector would result in a serial interval closest to that sampled serial interval. If a generation time or transmission interval was input, an equivalent approach is followed. This method ensures that transmission timings between individuals are biologically plausible and consistent with the chosen epidemiological assumptions. The simulator avoids producing negative intervals by rejecting any negative draws and optionally truncating the distribution at zero. Alternately, a user can choose to select infectors uniformly at random among all currently infectious individuals.

### Evolutionary model

Users can input their own reference strain from which all host genomic sequences will evolve, or simulate a random strain of fixed length. In either case, the user defines a mutation rate *µ* per nucleotide per unit time as the parameter for a Jukes-Cantor (JC) model of sequence evolution (equal substitution rates, no rate heterogeneity between sites). By default, all index cases are assigned the reference strain, but the package allows users to define multiple reference strains. Non-index cases are then simulated a sequence upon their time of infection, given their infector’s sequence and the length of time since their infector’s infection, under the JC model. The total number of mutations is Poisson-distributed with rate *µ · L ·* Δ*t*, where *µ* is the mutation rate, *L* is the genome length, and Δ*t* is the elapsed time since the parent strain’s realization.

### Outbreaks in heterogenously mixing populations

The ‘OOPidemic’ model also allows for simulation in heterogeneously mixing populations, through a structured/patch model. The simulation process is exactly as above, except the user defines multiple Group objects establishing multiple patches with potentially different numbers of initially infected and susceptible individuals, different reference strains, different epidemiological parameters e.g. lengths of infectiousness, and different transmission rates within and between groups. New cases are simulated per patch per time step, according to the force of infection coming into the patch. Infectors are chosen taking the relative force of infection from each patch into account. Although not used in the *R*_*t*_ study, this option is included in the R package allowing for more flexible outbreak simulations [17].

### Example outbreak simulations

In this section, we provide a series of example simulations using the ‘OOPidemic’ package, to demonstrate the consistency of our output simulations with the input parameters and highlight the information available to users.

All figures below result from simulations with genome length 29903 (as in SARS-CoV-2), and gamma distributed incubation periods, recovery periods, and serial intervals with mean and standard deviation (2, 0.9), (5.5, 1.2) and (3.5, 1.3) days, respectively. These are in line with many respiratory viruses e.g. influenza and human coronaviruses [24, 25]. For figure readability, we will simulate smaller outbreaks than those used in the main text ℛ_*t*_ experiments.

First, Figures 10 and 11 show outbreaks in homogeneously-mixing populations with transmission rate 0.4 among all individuals, and population sizes 200 and 2000, respectively. Figure 1 in the main text showed an equivalent outbreak in population size 1000. Even in the smallest outbreak, we observe that the simulated incubation, infectious and serial interval distributions match the input distributions well. Users have access to who-infected-whom (panel A), the compartment and incidence dynamics (panels B, C), individual time distributions (panels D, E, F) and genomic data for each case (panel G).

**Fig 10.**
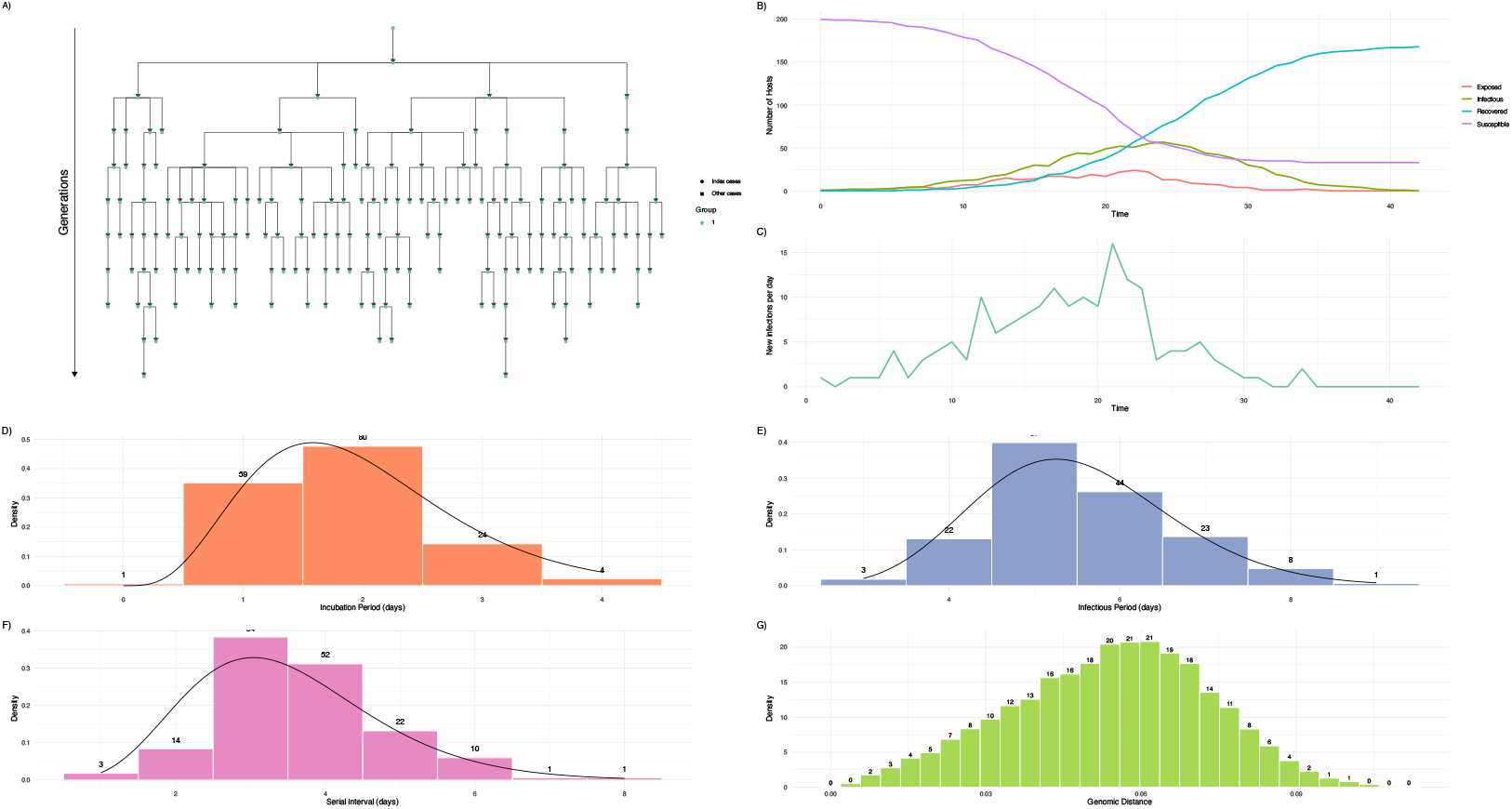
Simulated outbreak of size 168 in a heterogeneously mixing population with 200 hosts and 1 initial infector. Panels show the simulated (A) transmission tree, (B) compartment dynamics, (C) incidence curve, (D) distribution of incubation periods, (E) distribution of infectious periods, (F) distribution of serial intervals and (G) distribution of genomic distances between all possible pairs of infected hosts. In panels D, E, F, solid black lines show the gamma distributions simulated from.

**Fig 11.**
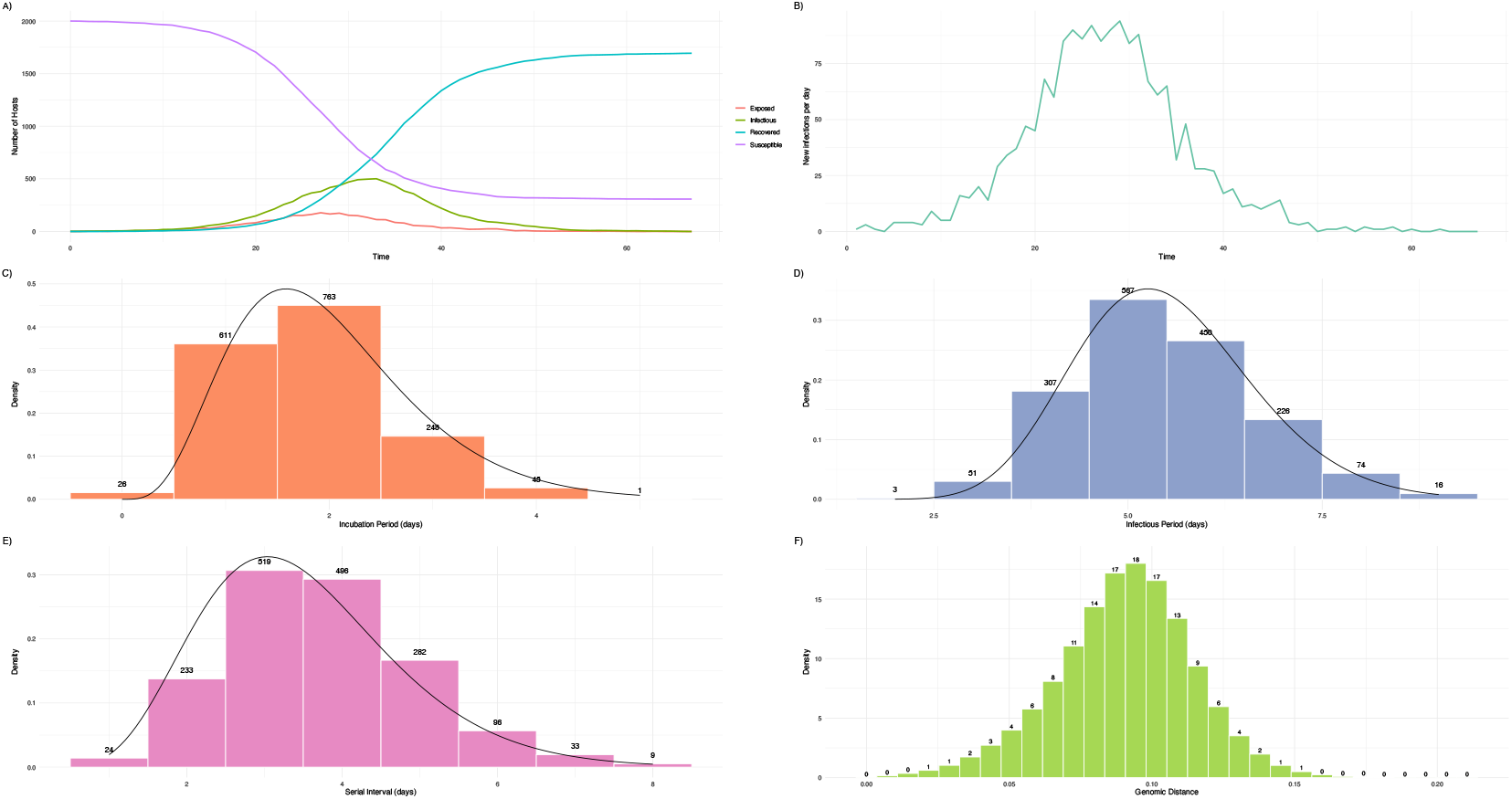
Simulated outbreak of size 1694 in a heterogeneously mixing population with 2000 hosts and 2 initial infectors. Panels show the simulated (A) compartment dynamics, (B) incidence curve, (C) distribution of incubation periods, (D) distribution of infectious periods, (E) distribution of serial intervals and (F) distribution of genomic distances between all possible pairs of infected hosts. In panels C, D, E, solid black lines show the gamma distributions simulated from.

Figures 12 through 17 demonstrate outbreaks simulated in heterogeneously-mixing populations using a patch model. Figures 12, 13, and 14 are structured with three groups of equal size (70, 300 and 900, respectively), with one initial infective per group, within-group transmission rate of 0.8 and between-group transmission rate of 0.1 among all groups. Figures 15, 16, and 17 show simulations structured with five groups of equal size (50, 200 and 400, respectively) and one initial infector per group. In Figure 15, we use a within-group transmission rate of 0.3 and between-group rate of 0.175. In Figures 16 and 17, we use a within-group transmission rate of 0.45 and between-group rate of 0.2625. That is, the 5-group simulations have a higher degree of between-group mixing than the 3-group. In smaller structured outbreaks (Figures 12 and 15), stochasticity can pull the observed distributions away from the expected, but as expected this is not the case in larger outbreaks. In addition to the information available in homogeneously-mixing outbreaks, users can track how many infections are occurring within and between-group (panels A).

**Fig 12.**
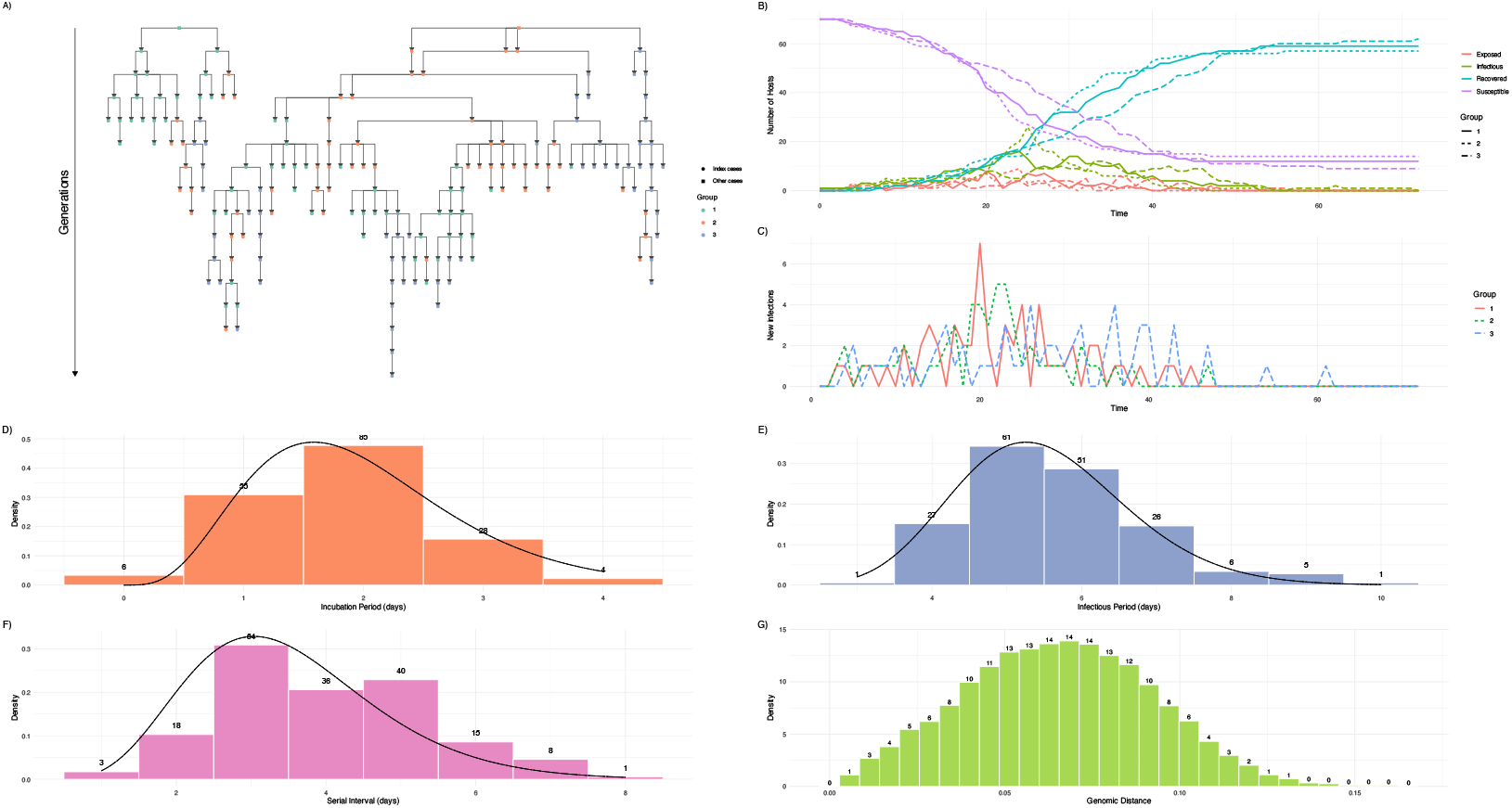
Simulated outbreak of size 178 in a 3-group population with 70 hosts per group and 1 initial infector per group. Panels show the simulated (A) transmission tree, (B) compartment dynamics, (C) incidence curve, (D) distribution of incubation periods, (E) distribution of infectious periods, (F) distribution of serial intervals and (G) distribution of genomic distances between all possible pairs of infected hosts. In panels D, E, F, solid black lines show the gamma distributions simulated from.

**Fig 13.**
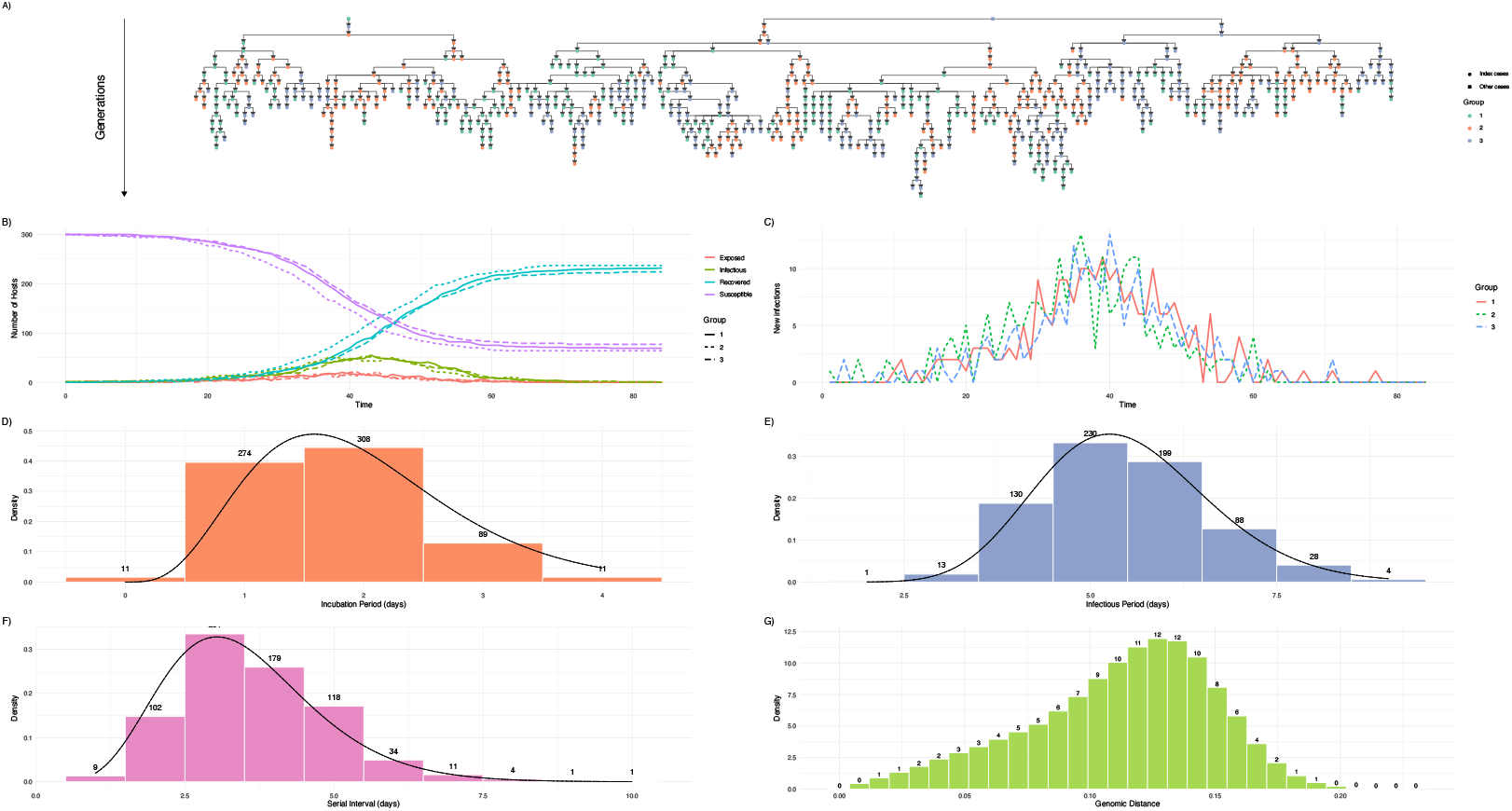
Simulated outbreak of size 693 in a 3-group population with 300 hosts per group and 1 initial infector per group. Panels show the simulated (A) transmission tree, (B) compartment dynamics, (C) incidence curve, (D) distribution of incubation periods, (E) distribution of infectious periods, (F) distribution of serial intervals and (G) distribution of genomic distances between all possible pairs of infected hosts. In panels D, E, F, solid black lines show the gamma distributions simulated from.

**Fig 14.**
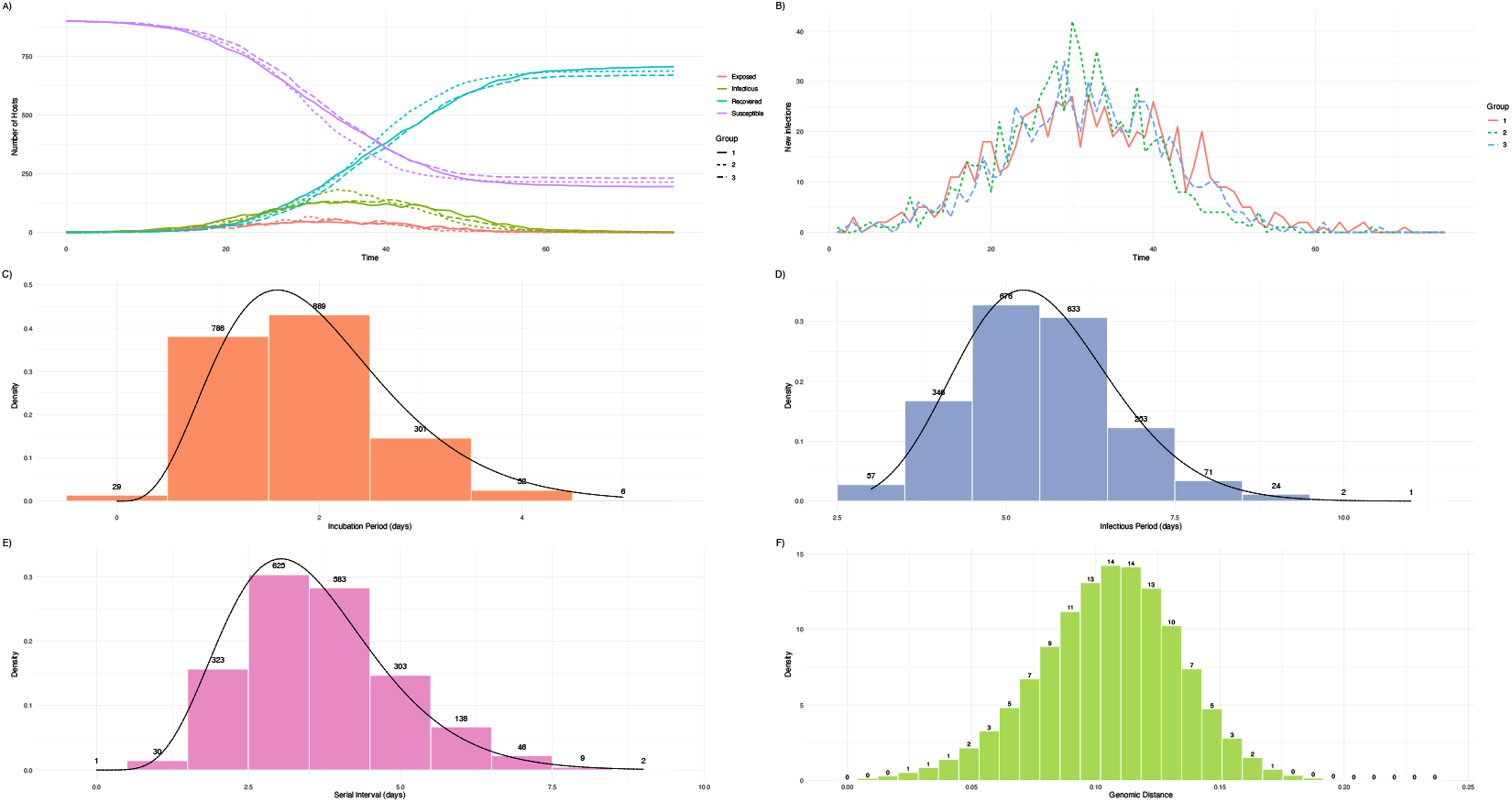
Simulated outbreak of size 2063 in a 3-group population with 900 hosts per group and 1 initial infector per group. Panels show the simulated (A) compartment dynamics, (B) incidence curve, (C) distribution of incubation periods, (D) distribution of infectious periods, (E) distribution of serial intervals and (F) distribution of genomic distances between all possible pairs of infected hosts. In panels C, D, E, solid black lines show the gamma distributions simulated from.

**Fig 15.**
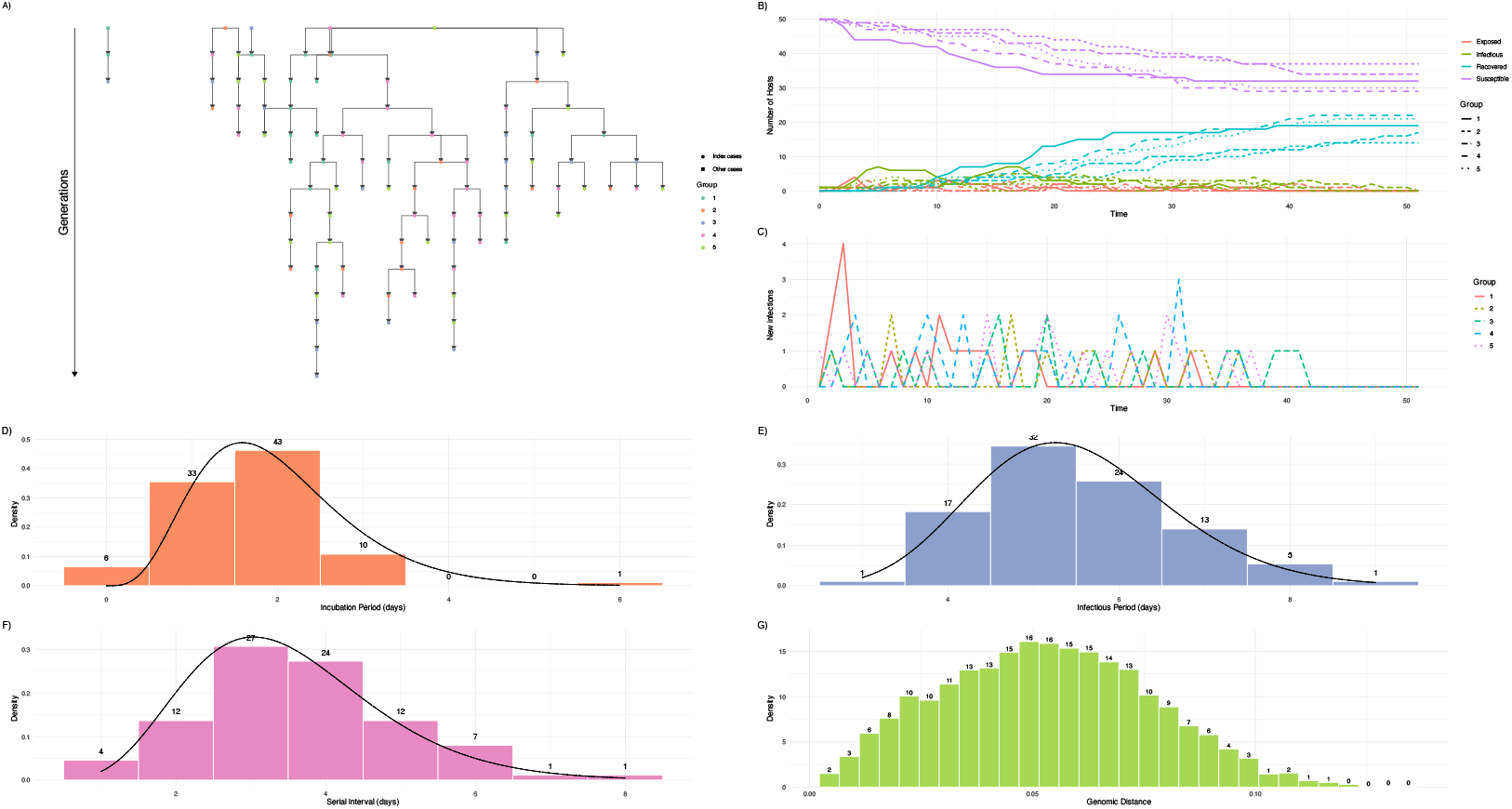
Simulated outbreak of size 93 in a 5-group population with 50 hosts per group and 1 initial infector per group. Panels show the simulated (A) transmission tree, (B) compartment dynamics, (C) incidence curve, (D) distribution of incubation periods, (E) distribution of infectious periods, (F) distribution of serial intervals and (G) distribution of genomic distances between all possible pairs of infected hosts. In panels D, E, F, solid black lines show the gamma distributions simulated from.

**Fig 16.**
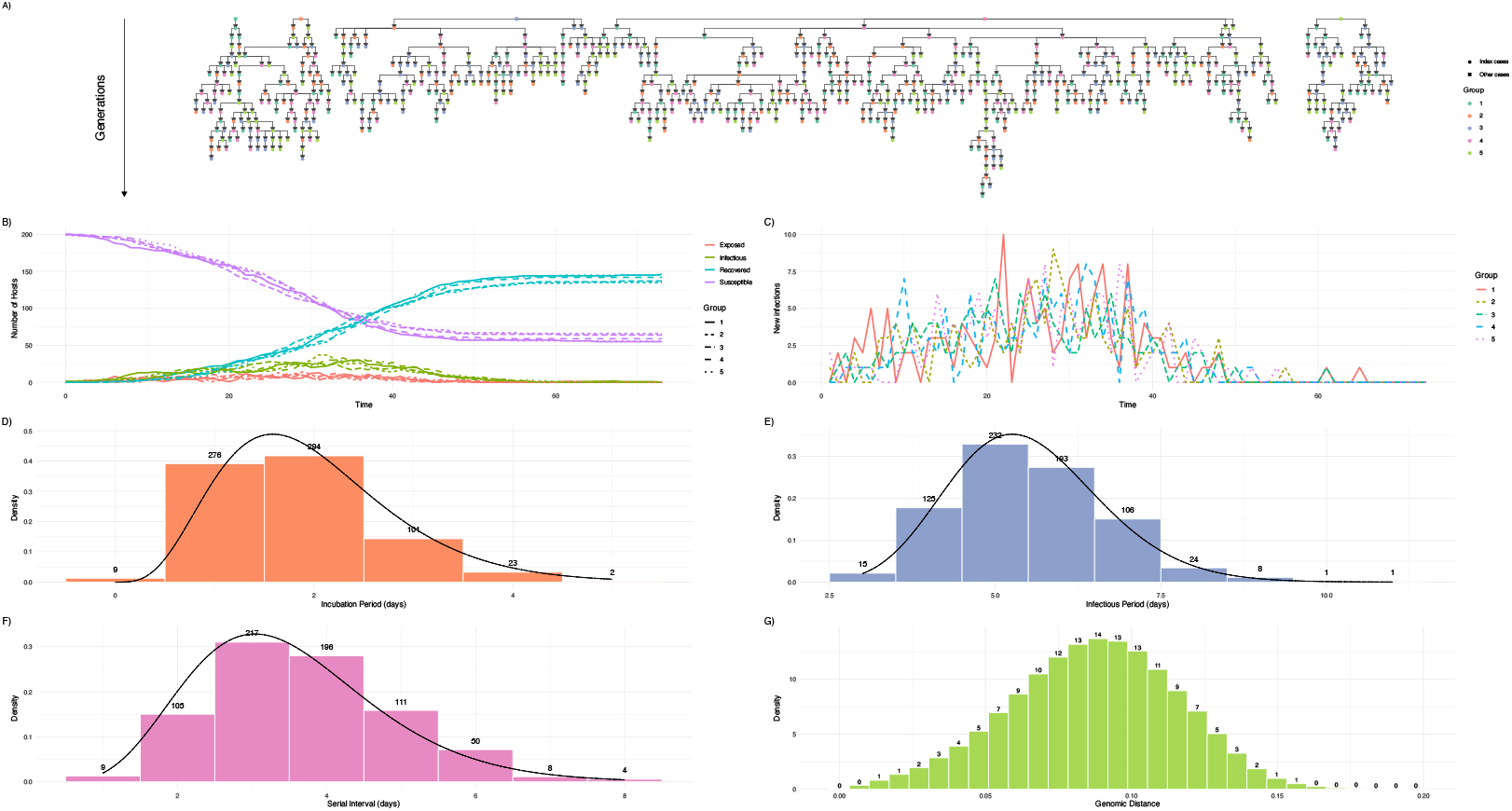
Simulated outbreak of size 705 in a 5-group population with 200 hosts per group and 1 initial infector per group. Panels show the simulated (A) transmission tree, (B) compartment dynamics, (C) incidence curve, (D) distribution of incubation periods, (E) distribution of infectious periods, (F) distribution of serial intervals and (G) distribution of genomic distances between all possible pairs of infected hosts. In panels D, E, F, solid black lines show the gamma distributions simulated from.

**Fig 17.**
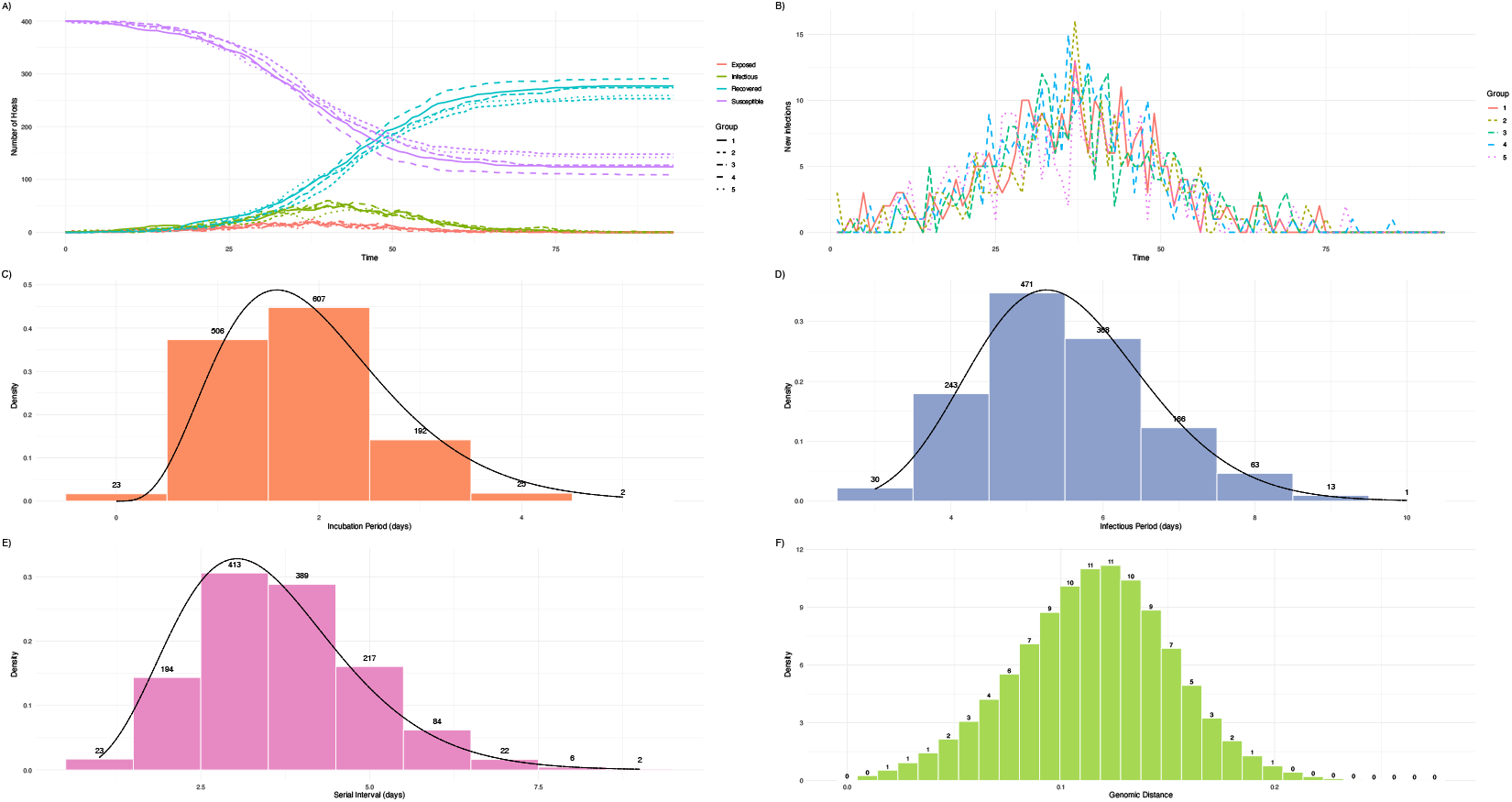
Simulated outbreak of size 1355 in a 5-group population with 400 hosts per group and 1 initial infector per group. Panels show the simulated (A) compartment dynamics, (B) incidence curve, (C) distribution of incubation periods, (D) distribution of infectious periods, (E) distribution of serial intervals and (F) distribution of genomic distances between all possible pairs of infected hosts. In panels C, D, E, solid black lines show the gamma distributions simulated from.

**Fig 18.**
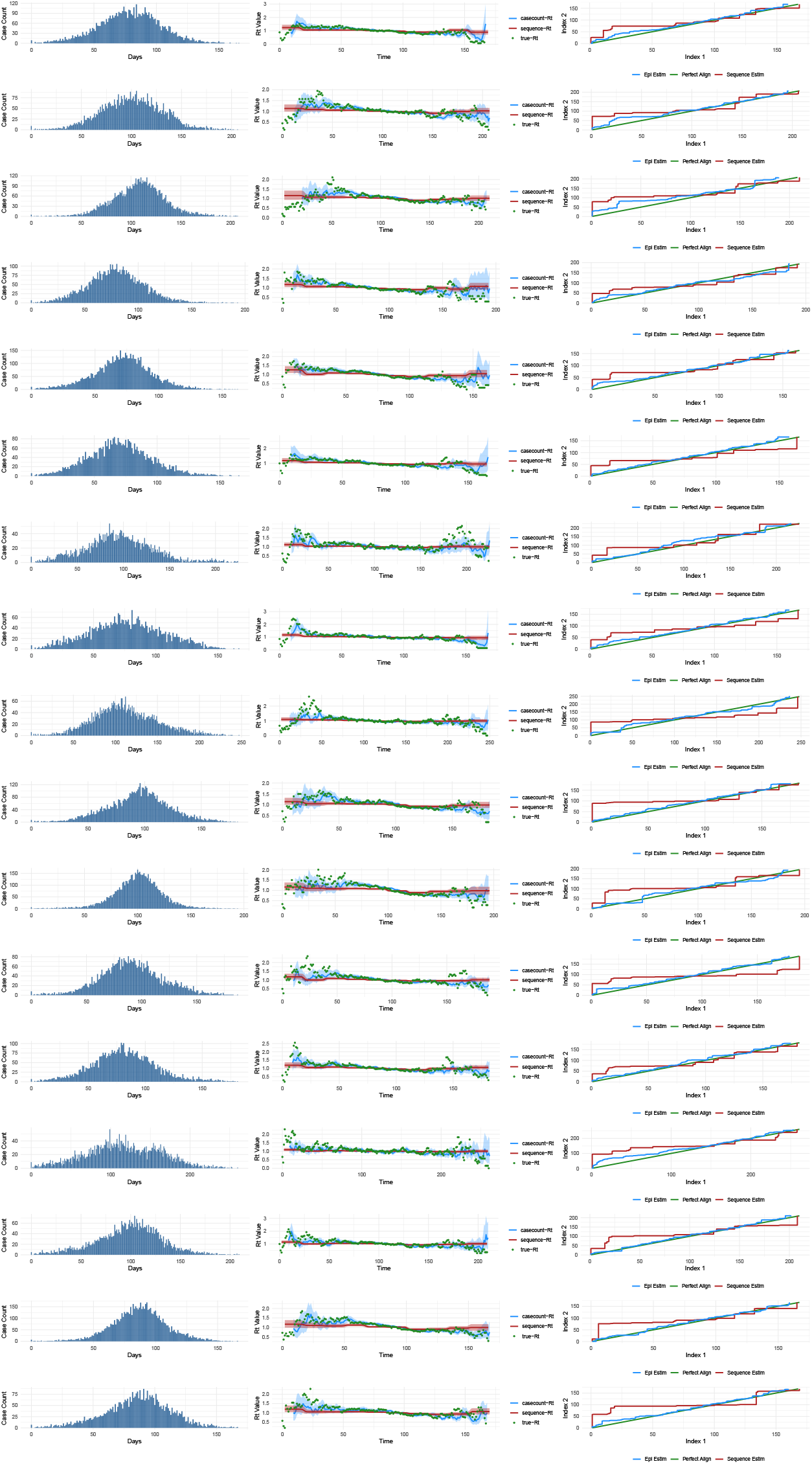
Outbreaks and comparison of accuracy of ℛ_*t*_ estimates from case count data and sequence data for the “optimum scenario”. The first column (bar plots) shows the incidence of cases per day over the course of the outbreak. The second column shows the actual estimates from the two data sources compared to the true ℛ_*t*_ estimate. The third column shows the alignment of each estimate with the true estimate. The green line represents the perfect alignment.

## Additional results

Additional results are presented here

**Fig 19.**
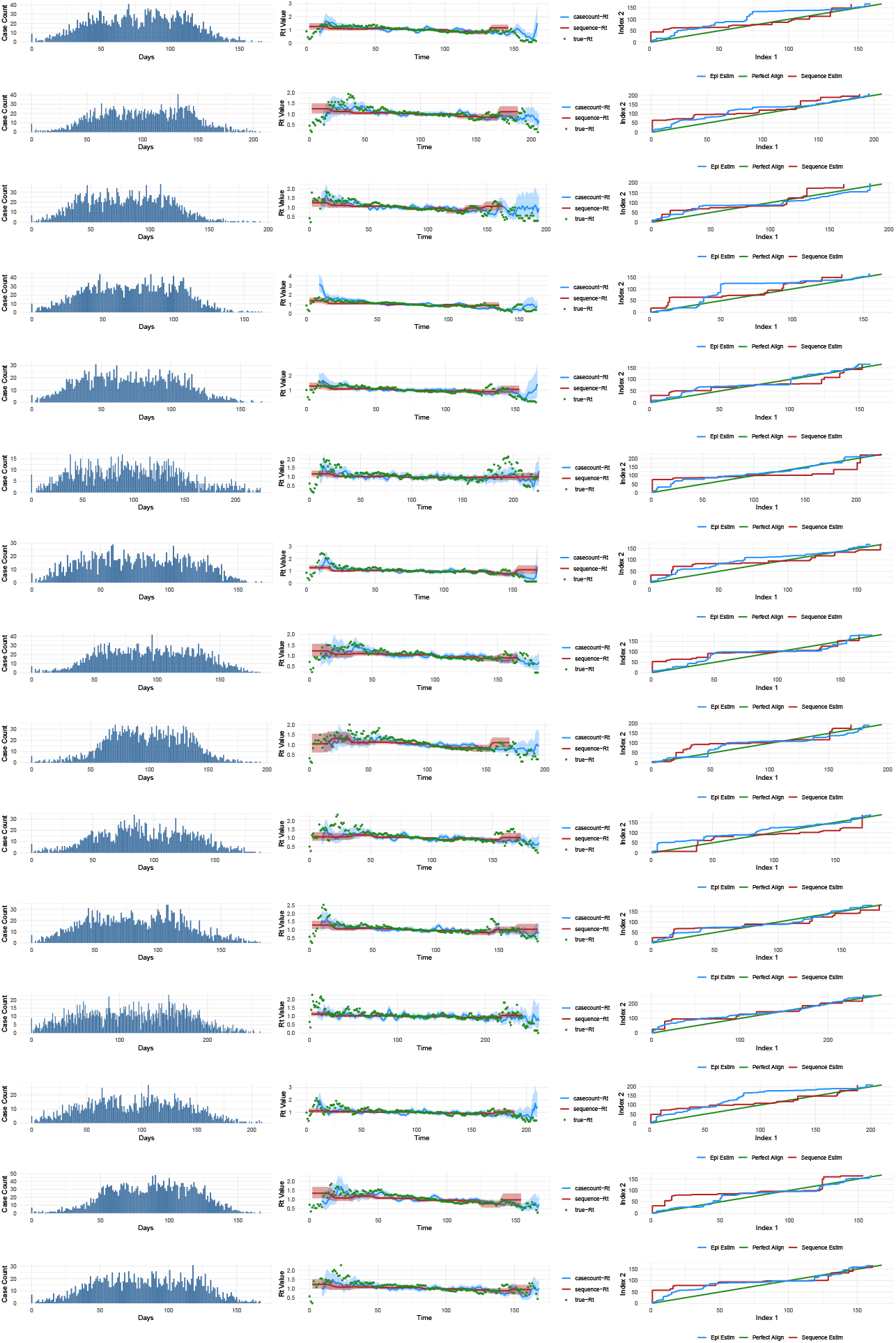
Outbreaks and comparison of the accuracy of ℛ_*t*_ estimates from case count data and sequence data for the 70% sampled scenario. The first column (bar plots) shows the incidence of cases per day over the course of the outbreak. The second column shows the actual estimates from the two data sources compared to the true ℛ_*t*_ estimate. The third column shows the alignment of each estimate with the true estimate. The green line represents perfect alignment.

**Fig 20.**
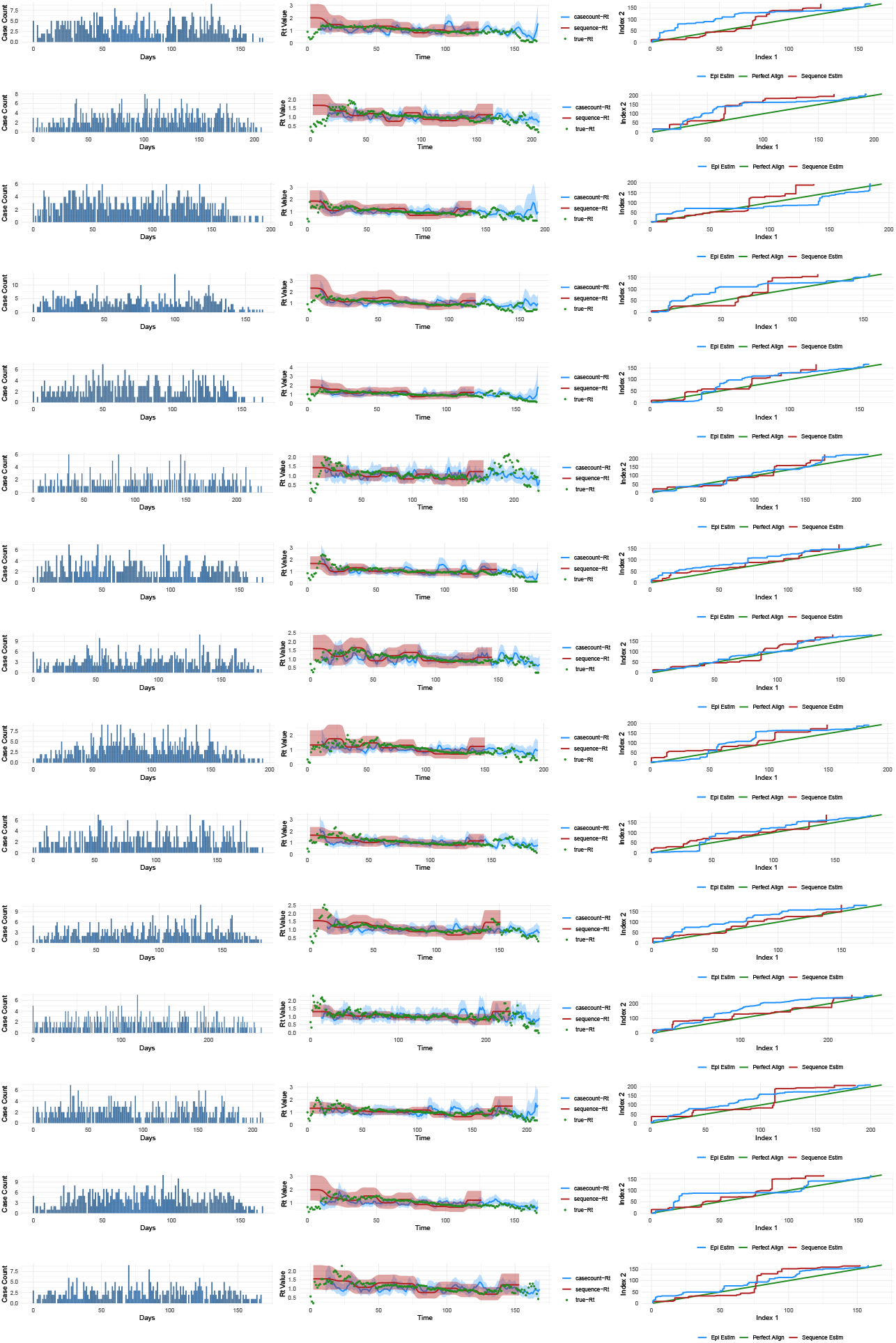
Outbreaks and comparison of the accuracy of ℛ_*t*_ estimates from case count data and sequence data for the 10% sampling scenario. The first column (bar plots) shows the daily incidence of cases over the course of the outbreak. The second column shows the estimates from the two data sources compared to the true ℛ_*t*_ estimate. The third column illustrates the alignment of each estimate with the true estimate. The green line represents perfect alignment.

**Fig 21.**
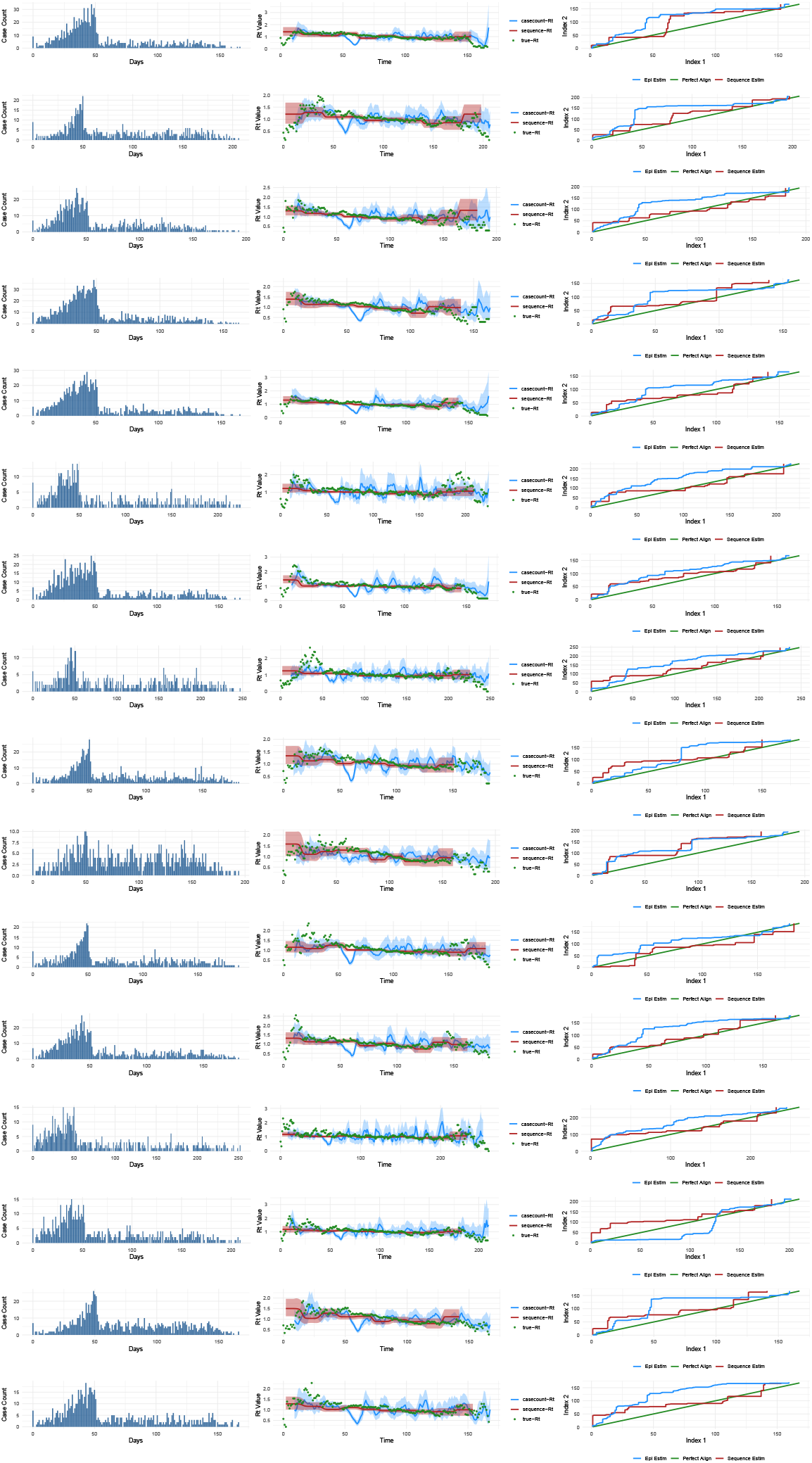
Outbreaks and comparison of the accuracy of ℛ_*t*_ estimates from case count data and sequence data for the shift from 70% to 10% sampling scenario. The first column (bar plots) illustrates the daily incidence of cases over the course of the outbreak. The second column displays the estimates from both data sources compared to the true ℛ_*t*_ estimate. The third column shows the alignment of each estimate with the true estimate. The green line indicates perfect alignment.

**Fig 22.**
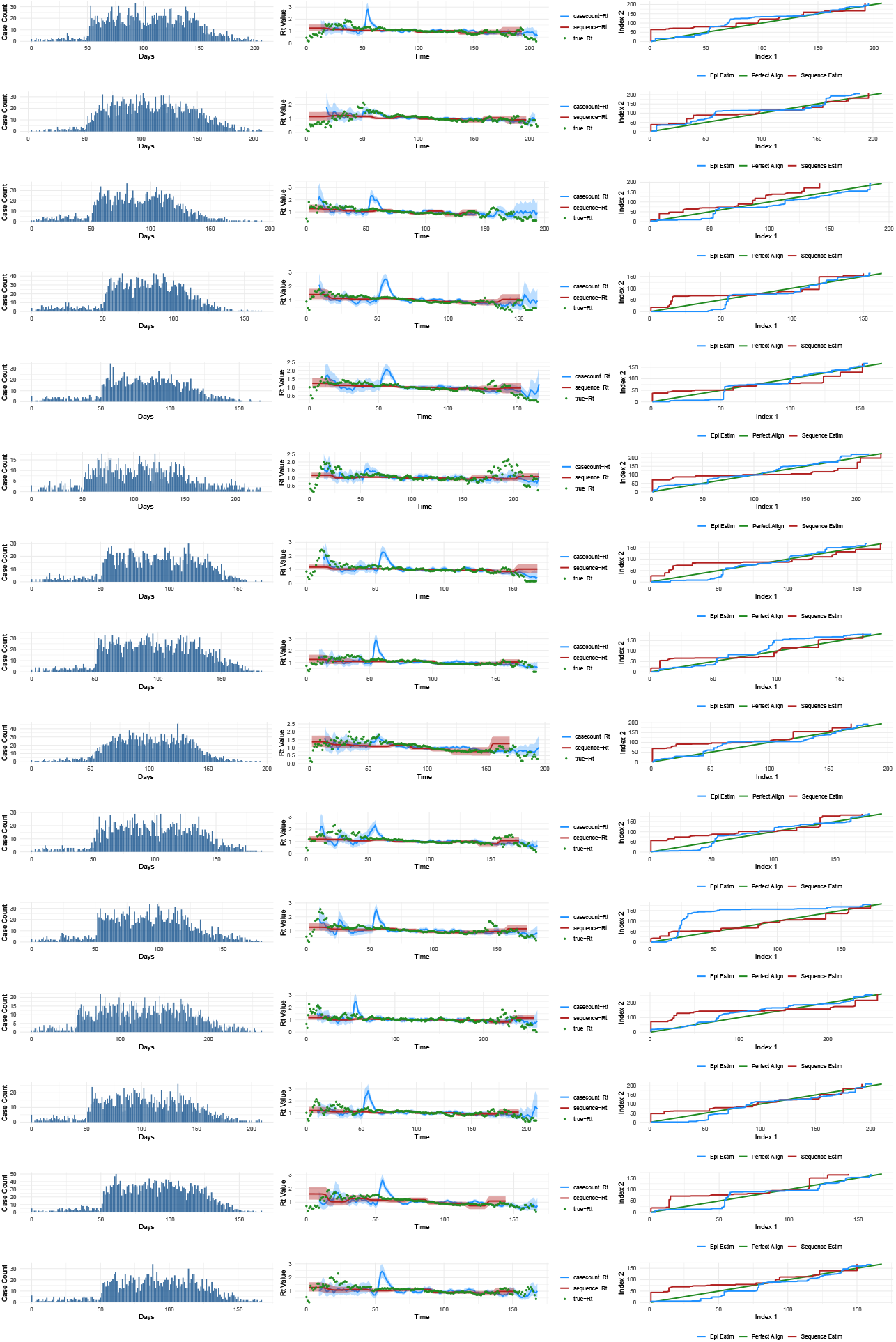
Outbreaks and comparison of the accuracy of ℛ_*t*_ estimates from case count data and sequence data for the transition from 10% to 70% sampling scenario. The first column (bar plots) depicts the daily incidence of cases throughout the outbreak. The second column presents the estimates from the two data sources in comparison to the true ℛ_*t*_ estimate. The third column demonstrates the alignment of each estimate with the true estimate. The green line represents perfect alignment.

**Fig 23.**
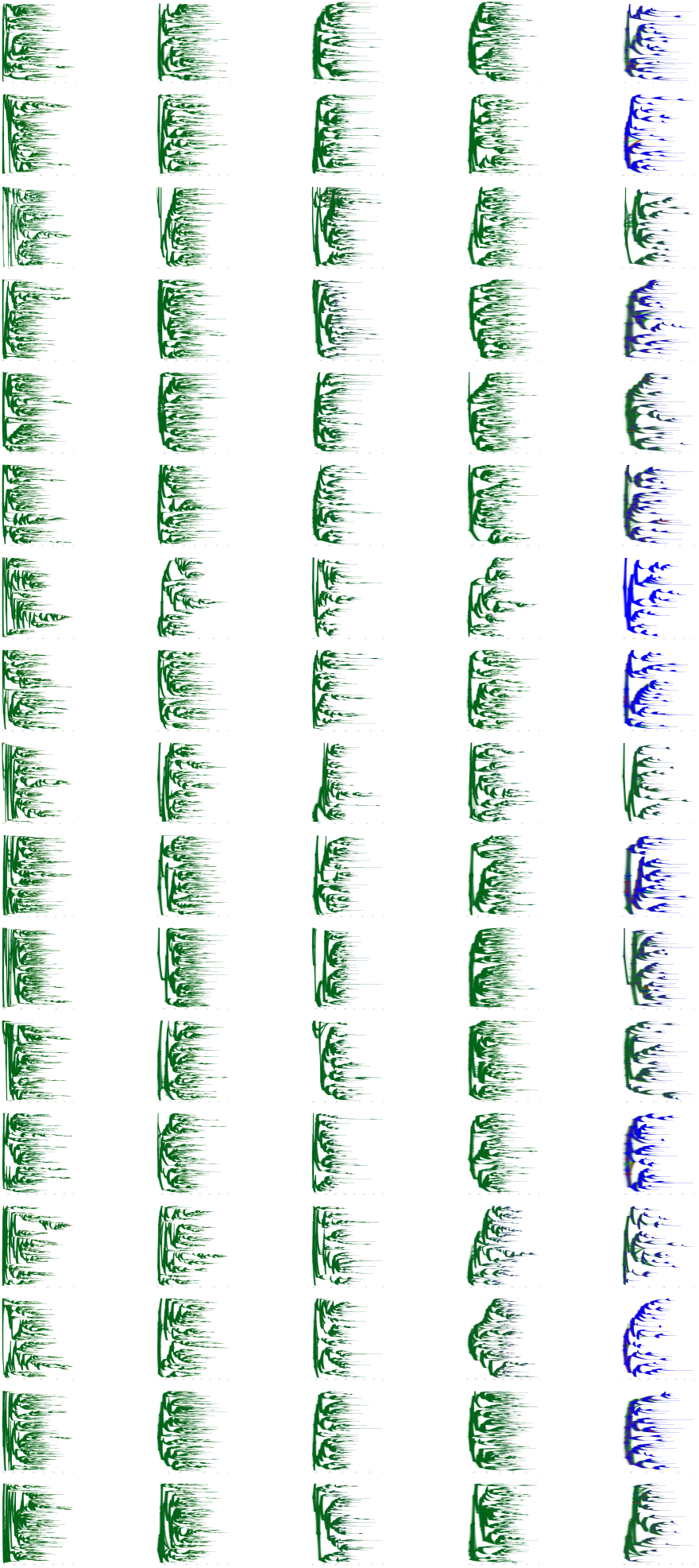
Comparison of tree posteriors (using densitree in R) from BEAST across different populations and sampling rates. Rows represent different populations, while columns correspond to different sampling regimens: optimum, 0.7, 0.7 to 0.1, 0.1 to 0.7, and 0.1. Trees shown in green indicate higher uncertainty due to a greater number of sequences, whereas trees in blue reflect lower uncertainty as they originate from the lowest sampling rate with fewer sequences.

## Notes

### Competing Interest Statement

The authors have declared no competing interest.

